# Dupilumab dampens mucosal type 2 response during acetylsalicylic acid challenge in N-ERD patients

**DOI:** 10.1101/2024.06.07.24308587

**Authors:** J. Eckl-Dorna, C. Morgenstern, K. Poglitsch, T. Arnoldner, K. Gangl, T. Bartosik, N.J. Campion, A. Tu, V. Stanek, S. Schneider, C. Bangert

## Abstract

**Rationale:** Non-steroidal anti-inflammatory drug-exacerbated respiratory disease (N-ERD) is characterized by the clinical triad of hypersensitivity to NSAIDs, nasal polyposis, and asthma. The cells and mediators causing acute symptoms when driving the hypersensitivity reaction to acetylsalicylic acid (ASA) ingestion, remain poorly defined.

**Objectives:** To investigate the dynamics of nasal mediators during ASA provocation in N-ERD patients before and twenty-four weeks after therapy with the IL-4 receptor alpha-blocking antibody dupilumab.

**Methods:** Nasal mucosal lining fluids of patients with N-ERD, chronic rhinosinusitis patients with nasal polyp (CRSwNP) and healthy disease controls were collected at selected time points up to two hours after ASA provocation. Analysis of thirty-three different inflammatory mediators as well as transcriptomic profiling was performed. In N-ERD patients, provocation was repeated after twenty-four weeks of dupilumab therapy.

**Measurements and Main Results:** Sixty minutes after provocation with ASA, N-ERD patients showed a significant increase in type 2 associated cytokines (i.e., TSLP, IL-5, and eotaxin-3) as compared to the other patient groups. This effect was diminished after twenty-four weeks of dupilumab therapy and was independent of the development of ASA tolerance. Transcriptomics revealed dampened upregulation of type 2 associated pathways genes (i.e., *AREG*) as well as enhanced downregulation of lipid (i.e., *ALOX15*) and peroxisome metabolisms (i.e., *NOS2*) at ASA provocation after dupilumab therapy.

**Conclusions:** Treatment with dupilumab leads to reduced nasal type 2 cytokine secretion and distinct changes in transcriptomic profile during ASA provocation, but changes in type 2 mediators show no association with tolerance development.

## Introduction

Non-steroidal anti-inflammatory drug (NSAID)-exacerbated respiratory disease (N-ERD) comprises the triad of asthma, chronic rhinosinusitis with nasal polyps (CRSwNP) and intolerance towards cyclooxygenase-1 (COX-1) inhibitors like acetylsalicylic acid (ASA)^1^. It is typically an adult-onset disease affecting about 7% of asthmatic and 10% of CRSwNP patients^2,3^. Two phases of the disease can be distinguished: the chronic phase, marked by a persistent burden of asthma and nasal polyposis due to baseline chronic inflammation of the respiratory tract, and the acute respiratory hypersensitivity reaction occurring only upon ingestion of COX-1 inhibitors^4^. The latter is characterized by multiple inflammatory symptoms like nasal congestion and coughing, often leading to wheezing, and sudden impairment in lung function within twenty to 120 minutes after oral administration of NSAIDs.

Whilst the role of an imbalance in the arachidonic acid pathway leading to overproduction of selected lipid mediators (e.g., cysteinyl leukotrienes (CysLTs)) in driving N-ERD is well established^5,6^, the impact of the immune system is not yet fully understood. Mast cells but also platelets, eosinophils, T helper 2 cells (Th2), B cells and innate lymphoid cells (ILCs) are thought to be key players in driving the accentuated type 2 response underlying N-ERD^7–10^. This is supported by the observation that the therapeutic monoclonal therapeutic antibody omalizumab, able to bind free IgE and thus to reduce IgE-mediated mast cell activation, not only leads to tolerance induction to ASA in 60% of N-ERD subjects, but also to reduced disease burden^11,12^.

Other biologics interfering with type 2 mediated pathways such as mepolizumab and dupilumab, targeting IL-5 and the IL-4 Receptor α (IL-4Rα), respectively, also decrease polyp size as measured by total polyp score (TPS) and – in the case of dupilumab – are also known to possibly induce ASA tolerance in a subset of patients^13,14^.

Although a reduction of type 2 mediators and associated cell subsets was observed in the chronic phase of the disease in patients undergoing therapy with the respective antibodies^15,16^, these studies have not addressed the impact of treatment on cytokine dynamics during acute hypersensitivity reactions. In this regards, data on immune kinetics following ASA challenges are scarce and indicate an increase in systemic proinflammatory IL-6^17^ and nasal eosinophils as well as ILC2 influx as early as one hour after provocation, but no increase in serum chemotactic proteins such Eotaxin-1^18^. Thus, despite the involvement of lipid mediators being firmly established, the contribution from other inflammatory mediators, especially from the type 2 pathway, to acute hypersensitivity reaction, remain enigmatic.

Here we aimed to investigate immune mediators and other factors driving the acute reaction to ASA in N-ERD patients using two complementary strategies: Firstly, we compared the kinetics of nasal immune mediator profiles during the first two hours following oral ASA provocation in N-ERD patients to those of CRSwNP and healthy controls (HC). Secondly, we investigated potential changes in the immune mediators and transcriptomic profile of N-ERD patients undergoing ASA challenge before and after dupilumab therapy to dissect the impact of IL-4Rα chain blockade on ASA-induced reactions.

## Methods

### Study design and patients

The eleven N-ERD patients included in this study were part of a cohort of thirty-one N-ERD patients undergoing therapy with dupilumab as reported previously^14^. All subjects signed informed consent and were recruited at the Department of Otorhinolaryngology and the Department of Dermatology at the Medical University Vienna, Austria. The study was approved by the local ethics committee (EK 1044/2020) and registered at EudraCT (2019-004889-18) and ClinicalTrials.gov (NCT04442256).

### Oral ASA challenge

Oral ASA challenges were conducted before and twenty-four weeks after start of dupilumab treatment as previously described^14^ (Fig. 1A). Positive reactions to ASA were defined by the onset of clinical symptoms and a drop (≥20%) in forced expiratory volume in one minute (FEV_1_). Briefly, increasing ASA doses (125 mg, 250 mg and 500 mg) were orally administered to N-ERD patients. Clinical indicators of upper and lower respiratory tract reactions including FEV_1_ and nasal mucosal lining fluids (nMLFs) were obtained before challenge (baseline) and at various timepoints (10, 20, 40, 60 and 120 minutes) up to two hours following provocation. Once symptoms in the upper or lower respiratory tract started, further nMLF samples and clinical parameters were taken at 10, 30, 60 and 120 minutes after reaction onset. The onset of symptoms was then defined as timepoint zero. Nasal mucosal cells for transcriptomic analysis were collected before and two hours after reaction onset in N-ERD patients. There was no post-reaction reading for NERD11 at provocation two and only ten minutes post-reaction reading for NERD02.

**Figure 1.**
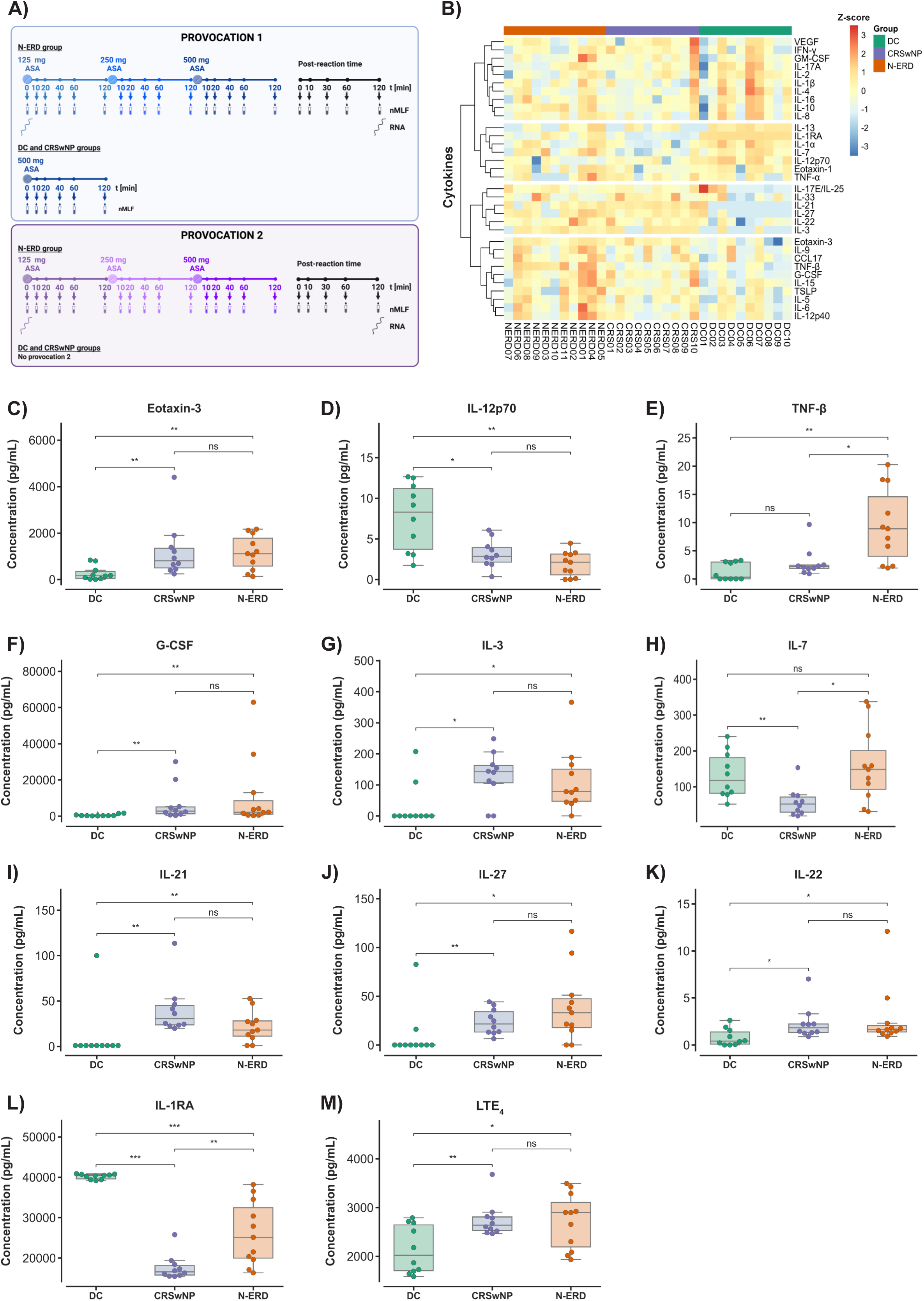
Differences in baseline nasal cytokine levels in disease controls (DC, green, n=10), chronic rhinosinusitis patients with nasal polyps (CRSwNP, purple, n=10) and non-steroidal anti-inflammatory drug (NSAID)-exacerbated respiratory disease (N-ERD, orange, n=11) patients. (A) Sampling timeline of oral ASA challenge at provocation 1 and 2. (B) Heatmap of nasal cytokine levels at baseline. Unsupervised clustering was performed on thirty-three Z-scaled nasal cytokines and chemokines using Euclidean distance and ward.2D linkage method. (C – M) Nasal levels of (C) eotaxin-3, (D) IL-12p70, (E) TNF-β, (F) G-CSF, (G) IL-3, (H) IL-7, (I) IL-21, (J) IL-27, (K) IL-22, (L) IL-1RA and, (M) Leukotriene E4 (LTE_4_) at baseline for the three patient groups (x-axes). Boxplots show distribution of absolute cytokine levels (y-axes, pg/mL) with median (horizontal line within box), quartiles (upper and lower box boundaries) and whiskers extending to 1.5 times the interquartile range. Individual patients are plotted as distinct data points on top. Group-wise comparisons were performed using the Mann-Whitney *U*-test for non-parametric cytokines and chemokines and the individual *t*-test for parametric leukotriene E4 data. Benjamini-Hochberg correction was applied for multiple testing. ns, not significant; \**p* < 0.05; \*\**p* < 0.01; \*\*\**p* < 0.001.

Disease controls and CRSwNP patients received the highest dose (500 mg) of oral ASA once and subsequently had nasal samples taken before challenge and at 10, 20, 40, 60 and 120 minutes.

For the methodology of the collection and processing of nasal samples, MSD assay (thirty-three inflammatory mediators), LTE_4_ and Tryptase assay, and RNA-sequencing please refer to supplemental material and methods.

### Statistical analysis and data processing

To assess if the data were normally distributed, histogram plotting and the Shapiro Wilk test were applied. The Kruskal-Wallis rank sum test was used to assess differences between multiple groups of non-parametric data. For *post-hoc* pairwise tests, Wilcoxon signed-rank and Mann-Whitney *U*-tests were applied for non-parametric dependent and independent comparisons, respectively. Differences in parametric variables were assessed by *t*-test. For comparisons of multiple binary outcome variables, Fisher’s exact test was used.

The Benjamini-Hochberg procedure was applied in multiple-testing to retrieve adjusted *p-*values. Significance was obtained at *p*-values smaller than 0.05.

All data processing and statistical analyses were performed with Python 3.9.6, R 4.3.1. and RStudio version 2023.09.0+463.

## Results

### N-ERD patients display a basal nasal mediator profile distinct from CRSwNP and healthy controls

In this exploratory study, ten patients with CRSwNP, eleven patients with N-ERD and ten disease control (DC) subjects were enrolled. Patient groups were well balanced except for DCs being younger than patients with polyposis (Table 1).

**Table 1.** Characteristics of study participants.

| Variable | DC, N = 10 | CRSwNP, N = 10 | N-ERD, N = 11 | *p*-value<sup>1</sup> |
| --- | --- | --- | --- | --- |
| **Sex, n (%)** |  |  |  | 0.88 |
| *female* | 3 (30%) | 2 (20%) | 4 (36%) |  |
| *male* | 7 (70%) | 8 (80%) | 7 (64%) |  |
| **Age, median (IQR)** | 32 (27, 41) | 44 (41, 50) | 47 (40, 57) | 0.028 |
| **Respiratory asthma, n (%)** | NA | 0 (0%) | 11 (100%) | <0.001 |
| **Allergy, n (%)** | NA | 4 (40%) | 9 (82%) | 0.080 |
| **Total polyp score, mean (SD)** | NA | 5.00 (1.70) | 4.00 (2.53) | 0.37 |

<sup>1</sup>Fisher's exact test; Kruskal-Wallis rank sum test
DC: Disease Control; CRSwNP: Chronic rhinosinusitis with nasal polyps; N-ERD: Non-steroidal anti-inflammatory drug (NSAID)-exacerbated respiratory disease; IQR: interquartile range; SD: standard deviation; NA: not applicable

When determining baseline levels of thirty-three different inflammatory mediators in nMLFs, we observed clearly distinct patterns of certain cytokines between the three patient groups (Fig. 1B-M, Fig. S1 and Table 2). CRSwNP and N-ERD patients showed increased mediator levels in two clusters containing alarmins (e.g., IL-33 and IL-17E/IL-25; Fig. 1B) and type 2 mediators (e.g., eotaxin-3, IL-9, CCL17; Fig. 1B), respectively. Interestingly, while most of the inflammatory mediators assessed were comparable between CRSwNP and N-ERD patients, we detected significant differences between these groups as compared to DC in the expression of the type 2 response-associated cytokine eotaxin-3 (Fig. 1C), differential levels of Th1-associated cytokines (IL-12p70, TNF-β; Fig. 1D and E) as well as an increase in cytokines involved in cell proliferation, differentiation and survival (G-CSF, IL-3, IL-7, IL-21; Fig. 1F-I), immune regulation (IL-27; Fig. J) and tissue repair (IL-22; Fig. 1K). Furthermore, these two patient groups demonstrated significantly reduced levels of the soluble IL-1 receptor antagonist (IL-1RA; Fig. 1L) and increased levels of leukotriene E4 (LTE_4_; Fig. 1M) in their nMLF.

**Table 2.** Baseline levels of nasal mediators in disease controls (DC), chronic rhinosinusitis patients with nasal polyps (CRSwNP) and non-steroidal anti-inflammatory drug (NSAID)-exacerbated respiratory disease (N-ERD).

| Mediator | DC, N = 10<sup>1</sup> | CRSwNP, N = 10<sup>1</sup> | N-ERD, N = 11<sup>1</sup> | p-value<sup>2</sup> |
| --- | --- | --- | --- | --- |
| IL-17E/IL-25 | 0.0 (0.0, 6.9) | 2.7 (1.9, 3.9) | 2.6 (2.2, 3.6) | 0.204 |
| IL-21 | **1.0 (1.0, 1.0)** | **30.8 (23.7, 45.1)** | **18.0 (11.3, 28.1)** | **0.002** |
| IL-22 | **0.4 (0.1, 1.4)** | **1.8 (1.3, 2.2)** | **1.6 (1.4, 2.1)** | **0.026** |
| IL-27 | **0.0 (0.0, 0.0)** | **21.5 (13.3, 34.2)** | **33.1 (17.8, 47.3)** | **0.008** |
| IL-33 | 9.5 (2.9, 28.5) | 44.2 (28.6, 55.2) | 7.3 (5.7, 43.1) | 0.065 |
| TSLP | 9.0 (3.7, 29.2) | 24.1 (11.8, 33.7) | 54.3 (9.2, 135.1) | 0.242 |
| Eotaxin-1 | 695.4 (396.9, 791.9) | 238.2 (228.2, 371.9) | 717.7 (210.8, 1,068.5) | 0.098 |
| Eotaxin-3 | **159.5 (45.9, 347.0)** | **806.0 (498.6, 1,343.9)** | **1,109.6 (575.7, 1,778.0)** | **0.003** |
| IL-12p70 | **8.3 (3.7, 11.2)** | **2.8 (2.1, 3.9)** | **2.1 (0.6, 3.1)** | **0.004** |
| IL-13 | 39.7 (29.2, 47.0) | 9.3 (3.0, 23.6) | 15.6 (3.0, 34.7) | 0.052 |
| IL-1RA | **40,439.3 (39,594.3, 40,667.4)** | **16,571.5 (15,784.5, 18,119.0)** | **25,121.7 (19,992.7, 32,451.4)** | **<0.001** |
| IL-3 | **0.0 (0.0, 0.0)** | **142.2 (106.7, 162.1)** | **78.3 (47.4, 150.8)** | **0.019** |
| IL-9 | 0.7 (0.0, 3.4) | 2.1 (1.1, 4.4) | 4.8 (1.9, 16.4) | 0.091 |
| CCL17 | 42.8 (25.1, 92.6) | 108.4 (35.0, 187.0) | 109.3 (76.5, 267.1) | 0.125 |
| TNF-α | 10.8 (2.0, 13.3) | 10.8 (5.9, 19.9) | 8.3 (0.0, 28.3) | 0.795 |
| VEGF | 2,774.2 (2,108.8, 6,714.3) | 4,157.9 (3,367.1, 6,413.9) | 4,177.0 (2,834.4, 5,106.5) | 0.759 |
| GM-CSF | 7.1 (4.8, 10.2) | 7.7 (5.8, 11.2) | 6.5 (4.6, 10.2) | 0.698 |
| IFN-γ | 9.8 (0.1, 74.7) | 21.0 (12.8, 35.4) | 18.2 (4.7, 22.2) | 0.323 |
| IL-10 | 3.8 (1.2, 19.6) | 2.2 (1.6, 2.9) | 7.9 (2.2, 12.6) | 0.245 |
| IL-17A | 35.6 (30.7, 127.8) | 30.3 (25.7, 40.0) | 26.0 (15.6, 46.5) | 0.481 |
| IL-1β | 112.1 (56.0, 353.8) | 43.8 (23.8, 123.2) | 135.3 (41.4, 186.8) | 0.298 |
| IL-2 | 63.2 (35.2, 115.4) | 38.9 (18.7, 60.3) | 27.0 (13.5, 63.4) | 0.250 |
| IL-4 | 5.6 (1.2, 17.5) | 1.5 (1.2, 3.0) | 2.4 (0.4, 5.4) | 0.305 |
| IL-5 | 9.2 (2.6, 88.3) | 32.7 (13.0, 97.7) | 176.8 (27.2, 465.2) | 0.059 |
| IL-6 | 102.2 (49.5, 259.7) | 78.3 (39.3, 103.2) | 203.3 (52.9, 760.5) | 0.376 |
| IL-8 | 10,810.2 (4,680.5, 28,945.8) | 6,651.5 (3,855.1, 14,037.7) | 12,443.5 (6,848.3, 15,548.8) | 0.543 |
| G-CSF | **239.4 (189.1, 557.3)** | **2,751.6 (1,301.5, 5,079.9)** | **2,194.6 (1,042.7, 8,524.9)** | **0.002** |
| IL-12p40 | 17.9 (12.6, 44.3) | 23.5 (16.5, 42.1) | 50.7 (16.4, 85.9) | 0.181 |
| IL-15 | 12.3 (8.7, 20.3) | 10.6 (9.4, 20.2) | 21.7 (17.1, 32.8) | 0.275 |
| IL-16 | 5,575.3 (1,597.5, 11,376.0) | 2,942.6 (1,771.9, 3,616.6) | 3,519.8 (2,413.5, 10,017.3) | 0.535 |
| IL-1α | **714.9 (366.3, 980.3)** | **53.5 (37.4, 232.6)** | **137.8 (61.4, 841.6)** | **0.001** |
| IL-7 | **117.6 (81.9, 181.1)** | **51.7 (29.0, 71.1)** | **148.2 (92.8, 200.6)** | **0.005** |
| TNF-β | **0.3 (0.0, 3.0)** | **2.2 (1.8, 2.4)** | **8.9 (4.0, 14.6)** | **0.003** |
| LTE<sub>4</sub> nasal | **2,022.1 (1,703.0, 2,644.4)** | **2,639.1 (2,528.1, 2,807.4)** | **2,895.3 (2,191.5, 3,104.6)** | **0.036** |
| LTE<sub>4</sub> urine | NA | 1,281.6 (958.7, 1,420.4) | 1,386.7 (1,211.7, 1,536.0) | 0.231 |
| Tryptase | 63.0 (14.4, 92.1) | 10.5 (5.4, 16.4) | 28.5 (9.7, 59.8) | 0.222 |

<sup>1</sup>Median (IQR)
<sup>2</sup>Kruskal-Wallis rank sum test

### Oral ASA provocation induces increased type 2 mediators after nasal provocation in N-ERD patients

Cytokine as well as tryptase and LTE_4_ levels were analysed at predefined timepoints after oral ASA provocation in patients with N-ERD, CRSwNP as well as DC (Fig. 1A). The median percentage change in cytokine concentration from baseline (=timepoint of ASA administration) for DC and CRSwNP and from reaction onset for N-ERD across 120 minutes was determined (Fig. 2A – H and Fig. S2A - AA). We observed distinct changes in mediator levels between the different groups and timepoints, peaking at sixty minutes after baseline for most mediators (Fig. 2A-H and Fig. S2A-AA). Interestingly, not only N-ERD, but also CRSwNP patients showed the highest LTE_4_ concentration 120 minutes after provocation thereby significantly differing from DC (Fig. 3A). The epithelial alarmin TSLP was significantly elevated in N-ERD patients as compared to CRSwNP and DC already after sixty minutes of symptom onset (Fig. 3B). This was accompanied by a peak in type 2-associated mediator levels such as IL-4 (Fig. S3A), IL-5 (Fig. 3C), CCL17 (Fig. 3D) and IL-10 (Fig. 3E) in N-ERD patients only. Cytokines associated with type 1 responses such as IL-12p40 (Fig. 3F), and TNF-β (Fig. 3G) were also specifically elevated in ASA-sensitive subjects after sixty minutes. Eotaxin-3 (Fig. 3H), GM-CSF (Fig. S3B), IFN-γ (Fig. S3C) and VEGF (Fig. S3D) concentrations were increased in both CRSwNP and N-ERD groups as compared to DCs after sixty minutes. The acute phase pro-inflammatory cytokine IL-6 started to rise ten minutes after aspirin provocation in N-ERD patients, peaked at sixty minutes, and showed a sharp decline thereafter (Fig. S2E and Fig. S3E).

**Figure 2.**
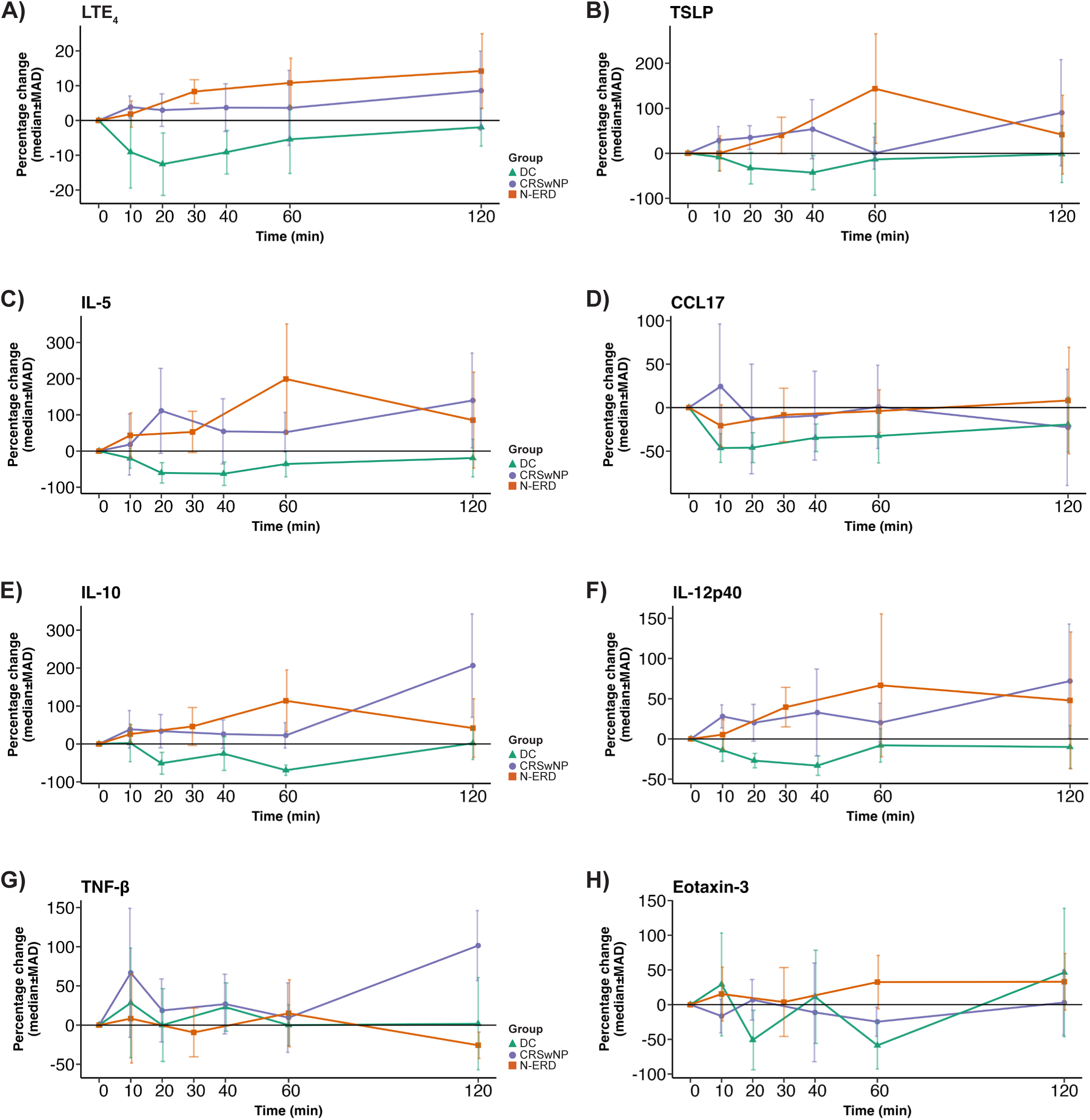
Percentage change in nasal mediator levels upon ASA provocation over time in disease controls (DC, green, n=10), chronic rhinosinusitis patients with nasal polyps (CRSwNP, purple, n=10) and non-steroidal anti-inflammatory drug (NSAID)-exacerbated respiratory disease (N-ERD, orange, n=11) patients. (A) – (H) Line graphs showing the median percentage change with median absolute deviation (MAD, y-axes) of cytokine levels of (A) LTE_4_, (B) TSLP, (C) IL-5, (D) CCL17, (E) IL-10, (F) IL-12p40, (G) TNF-β and (H) eotaxin-3 at indicated timepoints after ASA provocation in the three patient groups. Timepoint 0 was set at baseline for DC and CRSwNP and at reaction onset for N-ERD patients, respectively.

**Figure 3.**
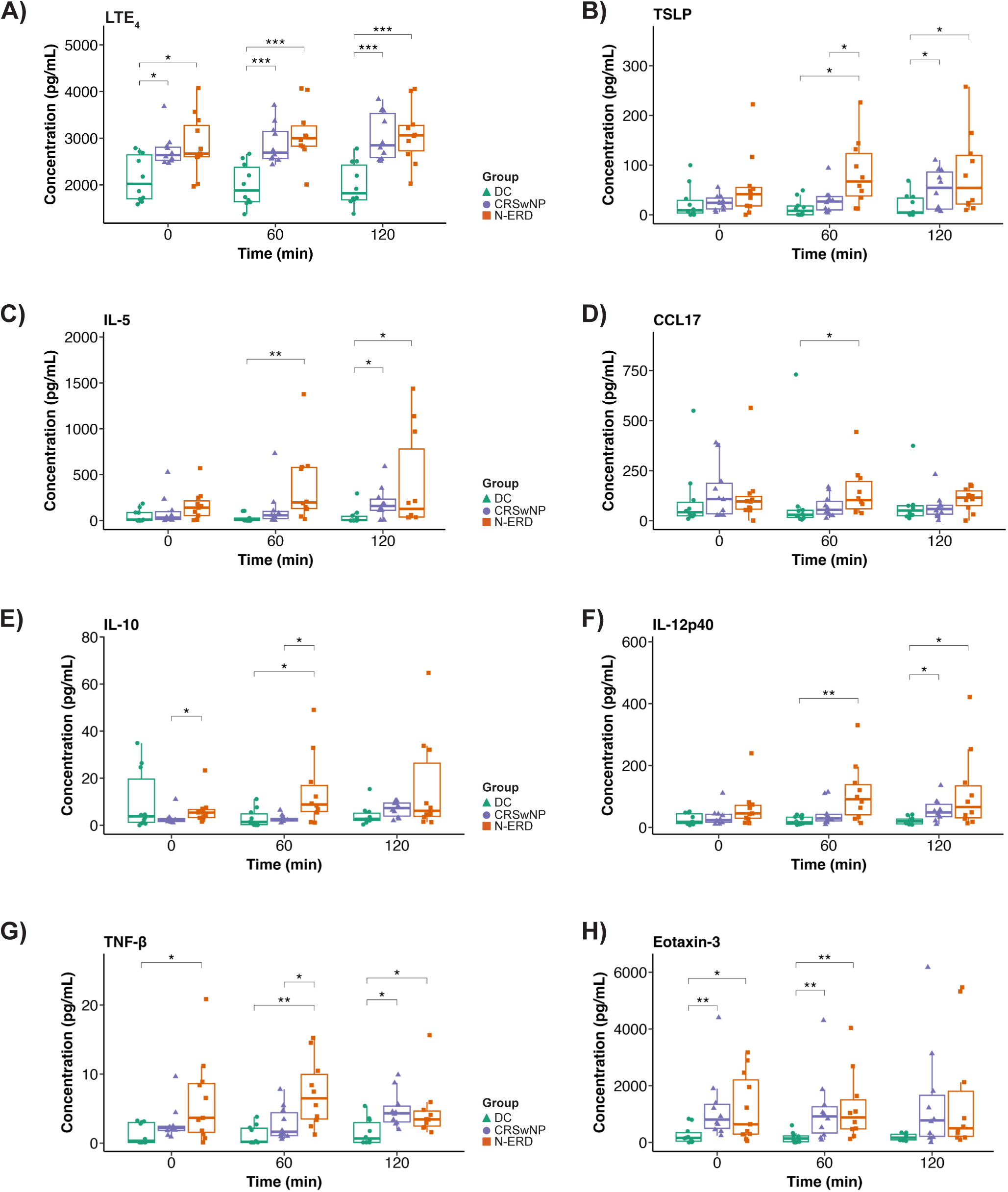
Changes in nasal mediator levels upon ASA provocation at selected time points in disease controls (DC, green, n=10), chronic rhinosinusitis patients with nasal polyps (CRSwNP, purple, n=10) and non-steroidal anti-inflammatory drug (NSAID)-exacerbated respiratory disease (N-ERD, orange, n=11) patients. (A) – (H) Boxplots showing absolute cytokine levels (y-axes, pg/mL) of (A) LTE_4_, (B) TSLP, (C) IL-5, (D) CCL17, (E) IL-10, (F) IL-12p40, (G) TNF-β and (H) eotaxin-3 at indicated timepoints after ASA provocation in the three patient groups (x-axes). Timepoint 0 was set at baseline for DC and CRSwNP and at reaction onset for N-ERD patients, respectively. Group-wise comparisons were performed using the Mann-Whitney *U*-test with Benjamini-Hochberg correction. \**p* < 0.05; \*\**p* < 0.01; \*\*\**p* < 0.001; non-significant not shown.

### Reduced secretion of type 2 inflammation-associated mediators in response to ASA provocation after 24 weeks of dupilumab therapy

Next, we were interested in assessing changes in ASA-induced nasal cytokine secretion in N-ERD patients before and after treatment with dupilumab. While baseline leukotriene levels were generally reduced after dupilumab treatment, as previously shown^14^, we were surprised to observe that N-ERD patients still expressed substantial amounts of LTE_4_ after the second ASA provocation (Fig. 4A and Fig. S5A). In contrast, secretion of TSLP in response to the ASA challenge was nearly abrogated after twenty-four weeks of dupilumab treatment (Fig. 4B and Fig. S5B) and the levels of other type 2 mediators such as IL-5 (Fig. 4C and Fig. S5C), CCL17 (Fig. 4D and Fig. S5D), eotaxin-3 (Fig. 4E and Fig. S5E) and IL-10 (Fig. 4F and Fig. S5F) were also significantly reduced. A similar pattern was observed for IL-12p40, TNF-β and VEGF in all patients (Fig. 4G-H and Fig. S5G-H, Fig. S4A and Fig. S5AH).

**Figure 4.**
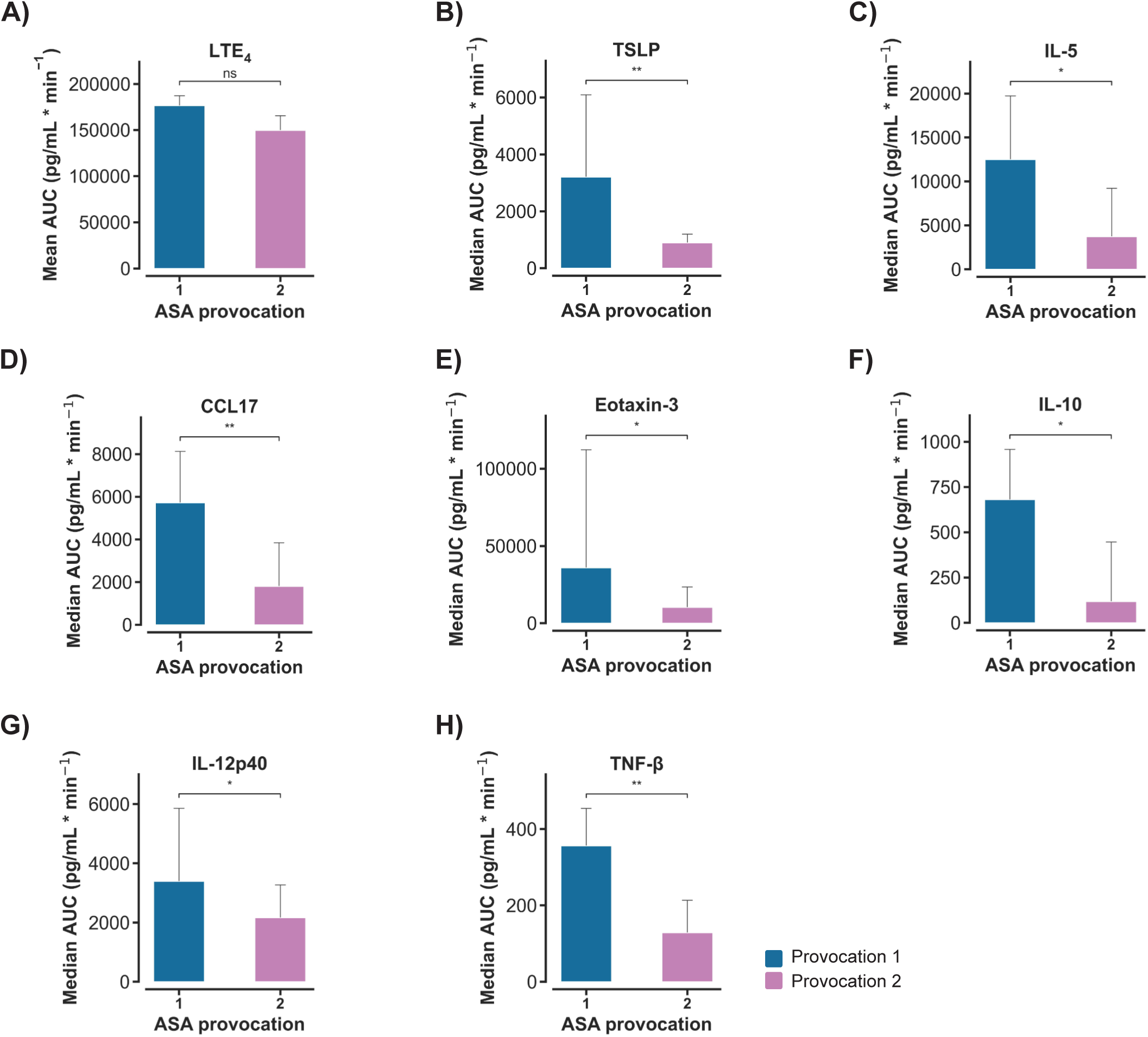
Changes in nasal mediator levels upon ASA provocation before (n=10, blue) and after 24 weeks (n=10, fuchsia) of dupilumab treatment in non-steroidal anti-inflammatory drug (NSAID)-exacerbated disease (N-ERD) patients. (A) – (H) Changes in (A) LTE_4_, (B) TSLP (C) IL-5, (D) CCL17, (E) eotaxin-3, (F) IL-10, (G) IL-12p40 and (H) TNF-β levels in N-ERD patients. Mean and median values of the area under the curve (AUC) from time point of reaction onset to sixty minutes post treatment of provocation 1 and 2 for LTE_4_ and cytokines, respectively. Error bars denote 95% confidence intervals. Differences were tested using *t*-test and Wilcoxon signed-rank test for LTE4 and cytokines, respectively. ns, not significant; \**p* < 0.05; \*\**p* < 0.01.

### Clinical improvement of N-ERD patients upon twenty-four weeks of dupilumab treatment

Immunological changes in type 2-associated molecules in our N-ERD cohort were accompanied by reduced symptom burden resulting in a significant decrease in TPS (Fig. 5A, *p*<0.01 (*p*=0.0048)) as well as improvement in olfactory perception as assessed by UPSIT (Fig. 5B, *p*<0.01 (*p*=0.0016)) after twenty-four weeks of dupilumab treatment^14^. This was associated with a decrease in general symptom burden as evaluated by SNOT-22 (Fig. 5C, *p*<0.01 (*p*=0.0027)) with nasal symptoms showing the greatest reduction (Fig. 5D, *p*<0.001, (*p*=0.00089)).

**Figure 5.**
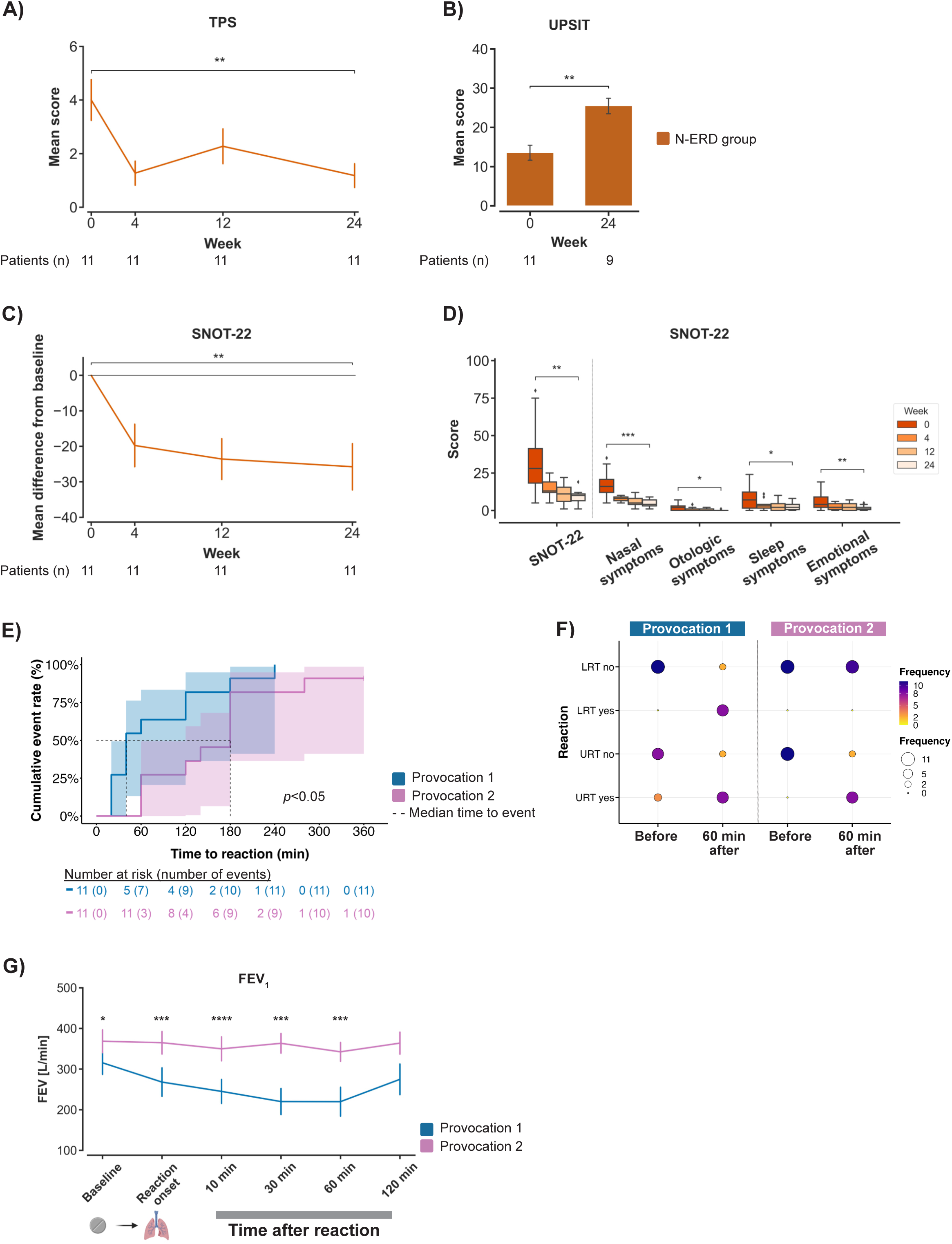
Changes in clinical parameters in non-steroidal anti-inflammatory drug (NSAID)-exacerbated disease (N-ERD) patients (n=11) treated with dupilumab for 24 weeks. (A) – (D) Change in (y-axes) (A) mean total polyp score (TPS), (B) University of Pennsylvania smell test (UPSIT) score, (C) sino-nasal outcome test-22 (SNOT-22) and (D) its subdomain scores (see methods) from week zero to week twenty-four as indicated (x-axis or (D) color coded). Error bars denote standard error of the mean. (E) Kaplan-Meier curves visualizing time to event (reaction to aspirin) data with 95% confidence intervals at provocation 1 (blue) and 2 (fuchsia). The log-rank test was applied to assess differences in the event rate between the two provocations. (F) Balloon plot showing the frequency of lower (LRT) and upper respiratory tract (URT) reactions to ASA or the absence thereof before and sixty minutes after reaction onset at provocation 1 and 2. (G) Levels of forced expiratory volume in one minute (FEV_1_) before and after reaction onset for provocation 1 and 2 as indicated (x-axis). Mean values of FEV_1_ are shown with standard error of the mean. Pairwise comparisons to assess differences between provocation 1 and 2 were done at each time point using the related *t*-test. \**p* < 0.05; \*\**p* < 0.01; \*\*\**p* < 0.001; \*\*\*\**p* < 0.0001; non-significant not shown.

Strikingly, the median time to symptom occurrence upon ASA provocation increased from provocation one to provocation two from approximately fifty minutes to 180 minutes (Fig. 5E). One patient was completely tolerant to ASA at the second provocation and four patients tolerated higher dosages, whilst six patients remained intolerant to ASA. Importantly, in contrast to the first provocation, all patients suffered only from upper but not from lower tract respiratory symptoms sixty minutes after ASA intake (Fig. 5F, Table S1). In accordance with this observation, patients had significantly higher FEV_1_ levels throughout the second provocation and showed a less pronounced drop in FEV_1_ after ASA intake (Fig. 5G). Patients developing ASA-tolerance (n=5) as compared to those remaining ASA-intolerant (n=6) did not show any difference by principal component analysis with regards to baseline cytokine levels (Fig. S6A-B) or in any of the modules identified at provocation one or two using the CytoMod software^19^ (Fig. S6C-F). Also, by comparing changes in cytokine levels in ASA-tolerant versus ASA-intolerant subjects between baseline and sixty minutes after the first and second provocation, we did not observe any significant differences for most mediators (Fig. S7A-AH) with the exemption of LTE_4_: subjects developing ASA tolerance had a significant increase in LTE_4_ concentrations during the first and a slight decrease during the second provocation, while those remaining intolerant showed an increase during both provocations (Fig S7AI).

### Transcriptomic signature of N-ERD patients after twenty-four weeks of dupilumab therapy

Changes at the transcriptomic level were assessed by comparing mRNA expression in nasal cells before and after ASA provocation, both at treatment start and after twenty-four weeks of dupilumab therapy. Significant differentially expressed genes (DEGs) were determined based on the following thresholds of log2 fold change (FC) and Benjamini-Hochberg adjusted *p*-value: |FC|>1.0, *p*<0.05. Among the top DEGs with the greatest log2 fold changes after the first ASA provocation were *CYP1B1* (−2.89 FC) encoding a member of the cytochrome P450 family involved in arachidonic acid metabolism^20^ and *IL5* (4.1 FC) (Fig. 6A). At the second provocation, the top downregulated genes were the cystatins *CST1* (−6.71 FC), *CST4* (−5.93 FC) and *CST2* (−5.62 FC) as well as *MUC5B* (−2.58 FC) amongst which *CST1* has been previously shown to be highly expressed in nasal polyps^21,22^ (Fig 6B). Compared to the first provocation, S*PINK 7*, a serine peptidase inhibitor attributed a role in barrier function and inflammatory responses^23^, as well as the fibroblast growth factor encoding protein *FGF5* were more upregulated at the second provocation, while proinflammatory mediators *CCL16,* and *IL11* as well as the antimicrobial peptide psoriasin *S100A7* showed less pronounced upregulation (Fig. 6C). *APOE*, which we previously observed to be downregulated in mast cells of N-ERD patients^10^ increased in expression during the second as compared to the first provocation (Fig. 6D). The genes downregulated before or during the second provocation included the calcium-activated chloride channel regulator (*CLCA1,* Fig. 6E), the 15-lipoxygenase (*ALOX15*, Fig. 6F) and the oxidative stress associated inducible nitric oxide synthetase *(NOS2,* Fig. 6G). Additional peroxisome associated genes (KEGG:04146) were also downregulated in individual patients at the second provocation, as depicted in Fig. 6H. Changes did not show any difference between ASA-tolerant and -intolerant subjects (Fig. S8A-D).

**Figure 6.**
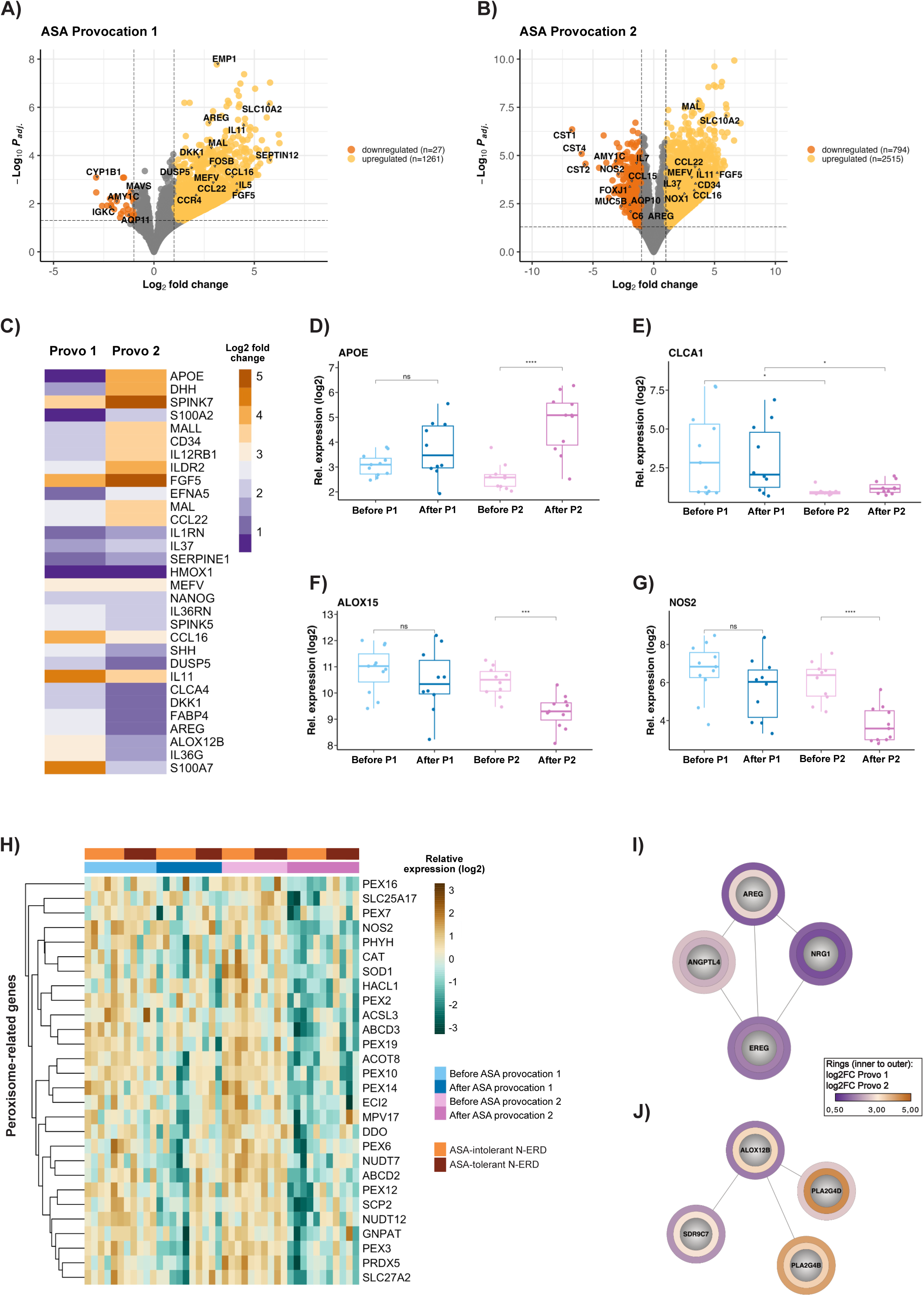
Nasal transcriptome signature of non-steroidal anti-inflammatory drug (NSAID)-exacerbated respiratory disease patients (N-ERD) before (n=11 before provocation 1 and n=10 after provocation 1) and 24 weeks after (n=10 before provocation 2 and n=11 after provocation 2) dupilumab therapy. (A) – (B) Differentially expressed genes at provocation 1 and 2. Volcano plots with up-and down-regulated genes upon ASA challenge at (A) provocation 1 and (B) 2. Dashed lines represent the cut-off values for adjusted *p*-values (*p*<0.05) and log2 fold changes (|FC|>1). (C) Heatmap with log2 fold changes of selected differential expressed genes between provocation 1 (Provo 1) and 2 (Provo 2). (D) – (G) Boxplots showing the relative expression (y-axes, log2) of (D) *APOE*, (E) *CLCA1*, (F) *ALOX15*, and (G) *NOS2* genes with median (horizontal line within box), quartiles (upper and lower box boundaries) and whiskers extending to 1.5 times the interquartile range before and after ASA challenge at provocation 1 (P1) and 2 (P2). Individual patients are plotted as distinct data points on top. Group-wise comparisons were done using a *t*-test. ns, not significant; \**p* < 0.05; \*\*\**p* < 0.001; \*\*\*\**p* < 0.0001. (H) Heatmap with relative log2 expression of peroxisome-related genes before and after ASA treatment at provocation 1 (blue) and 2 (fuchsia) and further stratified by ASA tolerance (red) and ASA intolerance (orange). (I) – (J) Clustered networks from Cytoscape analysis for (I) AREG and (J) ALOX12B related proteins. Grey filled circles represent the nodes, lines the network edges. Colored rings denote the log2 fold changes from differential expression analysis at provocation 1 (inner ring) and provocation 2 (outer ring).

Using Cytoscape network analysis, we found that amphiregulin (*AREG*), which is associated with type 2 immunity and directly modulates eosinophil responses^24^, was upregulated at both provocations albeit at lower levels after dupilumab therapy (Fig. 6I). The phospholipase A2 Group IVB encoding gene *PLA2G4B* was more upregulated at provocation two, whilst its isoenzyme *PLA2G4D* showed stronger expression at provocation one (Fig. 6J). Interestingly, enzymes of the 12-lipoxygenase pathway (*ALOX12B, SDR9C7,* Fig. 6J) also showed reduced upregulation at provocation two.

## Discussion

Here, we investigated - for the first time - cytokine dynamics in N-ERD patients during two hours following ASA provocation, not only as compared to ASA-challenged CRSwNP and DC, but importantly also before and after therapy with dupilumab. Sixty to 120 minutes after oral provocation with ASA, N-ERD patients not only showed a significant increase in type 2 associated cytokines (TSLP, IL-5, CCL17, eotaxin-3 and IL-10), but also in type 1 associated molecules such as IL-12p40 and TNF-β as compared to DC. Interestingly, ASA provocation resulted in increased eotaxin-3 and VEGF in both CRSwNP and N-ERD subjects after sixty minutes as compared to DC. Secretion of these cytokines upon ASA challenge was significantly reduced after twenty-four weeks of dupilumab therapy in all N-ERD patients. At the transcriptomic level, changes in the arachidonic acid pathway, type 2 associated pathways and the peroxisome metabolism were observed. While all eleven N-ERD patients improved clinically, only five developed partial or complete ASA tolerance. However, development of tolerance was not associated with specific cytokine patterns.

In accordance with previous data^9,25,26^, we observed an accentuated nasal type 2 cytokine pattern in CRSwNP and N-ERD patients at baseline, with the latter showing the highest levels of type 2 cytokines. Additionally, cytokines such as VEGF or eotaxin-3 were upregulated in both groups sixty minutes after ASA provocation. These observations corroborate the current notion that cytokine patterns underlying N-ERD and eosinophilic nasal polyps are identical, but that sheer quantity of these cytokines is the decisive factor determining the severity of disease symptoms^6^.

The pathophysiological mechanism underlying N-ERD involves an imbalance in arachidonic acid metabolism with increased levels of CysLTs. The acute reaction after NSAID intake is thought to be partly induced by blocking COX-1-mediated constitutive prostaglandin E2 secretion, which under stable conditions inhibits eosinophil and mast cell activation by ASA ingestion. However, recent evidence suggests additional, so far unidentified stimuli to be involved in eosinophil and mast cell activation with both IL-4 and IFN-γ playing an important role in mediating the phenotype of N-ERD^27^. Platelets, which are more activated in N-ERD patients and contribute to the chronic disease^28,29^, show no additional changes during ASA provocation testing^30^. Recent work suggests that innate lymphoid cells type 2 (ILC2) are implicated in acute reactions as they can be activated by PGD2, are found in increased numbers in nasal scrapings after ASA administration^31^ and could lead to tissue eosinophilia by producing large amounts of IL-5 shortly after ASA provocation^18^. Furthermore, the epithelial barrier dysfunction promoting cytokines IL-6 and oncostatin M show elevated levels in N-ERD patients as compared to healthy controls^32^. Our findings of increased TSLP levels, IL-4 and IL-5 as well as high eotaxin-3 levels sixty minutes after ingestion of ASA support the hypothesis of type 2 associated cell recruitment, including ILC2 as well as eosinophils during the acute reaction^33^.

As dupilumab inhibits both IL-4 and IL-13 mediated signaling, it can impact a variety of mechanisms involved in N-ERD pathophysiology ranging from Th2 cell differentiation, ILC2 expansion, mast cell activation to airway smooth muscle cell proliferation^34^. We and others have previously demonstrated that dupilumab leads to a marked downregulation of type 2 mediators in serum and nasal secretions of N-ERD patients over time^14,16,35^. However, here we show for the first time that the same trend is also demonstrable during ASA provocation after twenty-four weeks of dupilumab therapy. Interestingly, patients becoming tolerant to ASA during dupilumab treatment showed a similar cytokine pattern - excluding LTE_4_ - during the second provocation, compared to those remaining intolerant. Nevertheless, these findings may indicate that although activation of type 2 immunity plays a central role in driving the chronic phase of N-ERD and is also involved during ASA ingestion, the symptoms during the acute reaction in a majority of patients may be driven by additional, type 2 mediator-independent mast cell and eosinophil activation such as IgE. The fact that we observed attenuated LTE_4_ secretion during the second provocation in ASA-tolerant, but not intolerant subjects, corroborates this hypothesis. In line, the IgE targeting monoclonal antibody omalizumab^11,12^, induces complete tolerance to ASA in 56% of patients after twenty-four weeks of therapy, as compared to only 23% in patients treated with dupilumab^14^. This, together with the fact that IgE positive plasma cells as well as total IgE levels are elevated in polyps of N-ERD patients^36^, argues for a chronic IgE-induced activation of mast cells as an important factor in supporting acute reactions in N-ERD.

In N-ERD patients both the COX- and LOX-pathways are known to be dysregulated, leading to an increased production of cysteinyl leukotrienes (CysLTs). Previously, *ALOX15* was found to be upregulated in nasal polys and levels of *ALOX15* in epithelial cells of N-ERD patients correlate with disease severity^10,37^. Interestingly, we observed that dupilumab treatment results in reduced expression levels of genes encoding enzymes of both the 15-lipoxygenase (*ALOX15*) and the 12-lipoxygenase (*ALOX12B, SDR9C7*) pathways at the second provocation as compared to the first. This is in agreement with previous studies which found *ALOX15* to be downregulated after dupilumab treatment.^38^ At the same time, *CYP1B1* encoding cytochrome P450, which is responsible for metabolizing arachidonic acid to 20-hydroxyeicosatetraenoic acid and has been shown to lead to relaxation of airways^20,39^, was strongly downregulated during provocation one but not provocation two. This could indicate that long-term treatment with the anti-IL-4Rα inhibitor dupilumab may ameliorate the imbalance in the arachidonic acid pathway. Additionally, various genes being implicated in a role in CRSwNP and especially N-ERD pathology show reduced expression levels after ASA provocation upon twenty-four weeks of dupilumab treatment: i) *CST1* and other members of the cystatin family that play a role in amplifying type 2 inflammation^21^ and correlate with disease status and polyp burden^22,40^ ii) *MUC5B*^41^ and *CLCA1*^42^, which are both overexpressed in CRSwNP and play an important role in the regulation of mucus production and electrolyte transport iii) *NOS2,* the oxidative stress-associated nitric oxide synthetase, an important biomarker of type 2-high asthma and highly expressed in CRSwNP^43^. Together with *NOS2*, we found twenty-seven more downregulated DEGs that are associated with the peroxisome in gene set enrichment analysis (Fig. 6H, Table S2). On the other hand, transcription of the *APOE* gene encoding apolipoprotein, which ameliorates airway hyperreactivity in asthma ^44^ and is known to be downregulated in N-ERD patients^10^, is increased during the second provocation. Thus, dupilumab therapy induces a variety of changes in the transcriptome leading to alleviation of airway hyperresponsiveness and oxidative stress improving acute reaction symptoms to ASA challenge.

Limitations of this study include the small sample size of ten patients per group as well as the lack of CRSwNP and DC groups at provocation two due to the nature of this pilot trial. Furthermore, although our panel of cytokines was carefully selected, it is still only a fraction of globally available mediators.

In summary, our work provides first insights into the type 2 immunity associated processes governing the response to ASA provocation with and without dupilumab therapy. Our results warrant investigations in a larger cohort of patients undergoing long-term therapy with dupilumab using large-scale proteomics as well as single cell analysis to further delineate the factors involved in acute reactions to ASA.

## Data Availability

Data has been deposited at the European Genome-phenome Archive (EGA), which is hosted by the EBI and the CRG, under accession number EGAD50000000565. Further information about EGA can be found at https://ega-archive.org and "The European Genome-phenome Archive of human data consented for biomedical research".

## Acknowledgement

Figure 1A was created with BioRender.com.

## CLINICAL IMPACT

In this study, the impact of dupilumab on inflammatory mediator release and transcriptional changes during ASA provocation in patients suffering from NSAID-exacerbated respiratory disease was assessed for the first time.

## AUTHOR CONTRIBUTION STATEMENT

SS, CB, TA and JED designed the study, wrote the study protocol and gained ethical approval for the study. SS, KP, TA, KG, CS, TB, CB and NJC recruited all the patients and carried out the study. AT and VS performed the experiments. SS, CM, CB and JED performed the analysis. JED, CM, SS, and CB wrote the manuscript. All authors critically revised the manuscript together.

## Abbreviations

ASA: acetylsalicylic acid
CRSwNP: Chronic rhinosinusitis with nasal polyps
COX-1: Cyclooxygenase-1
CysLTs: Cysteinyl leukotrienes
FEV_1_: Forced expiratory volume in one minute
N-ERD: Non-steroidal anti-inflammatory drug
(NSAID): exacerbated respiratory disease
TPS: Total Polyp Score
ILC2: Type 2 innate lymphoid cells
nMLF: Nasal mucosal lining fluid
UPSIT: University of Pennsylvania Smell Identification Test
SNOT-22: Sino-nasal Outcome Test

