## supplemental material for "Dupilumab dampens mucosal type 2 response during acetylsalicylic acid challenge in N-ERD patients"

Sven Schneider, MD

Department of Otorhinolaryngology

Medical University of Vienna, General Hospital of Vienna

Waehringer Guertel 18-20

A-1090 Vienna, Austria

#### SUPPLEMENTARY METHODS

*Assessment of clinical endpoints (TPS, UPSIT, SNOT-22)*

The total poly score (TPS) was assessed endoscopically and ranges from 0 (no polyp) to 4 (large polyps) for each nostril. Changes in the mean TPS score between baseline and dedicated study time points in N-ERD patients were determined by paired *t*-test.

The forty item University of Pennsylvania Smell Identification Test (UPSIT) was used to measure the individual's ability to detect odors at baseline and end of study. The change in mean UPSIT score was assessed by individual *t*-test.
Twenty-two items of the Sino-nasal Outcome Test (SNOT-22)[1] test were analysed as total sum as well as split into the four domains (nasal symptoms: NS01-NS09, otologic symptoms: OS1-OS3, sleep symptoms: SS01-SS05, emotional symptoms: ES01-ES05). Missing data were imputed as reported previously[2]. Briefly, data from questionnaires were included in the analysis if they were greater than 80% complete and contained at least two patient visits. Missing values were imputed using the mean of the two closest visits. Item-total correlations were assessed by calculating Cronbach's Alpha and Spearman-Brown Coefficient using the Python library *psython*.

*Collection and processing of nasal samples*

Nasosorptions were transported on ice from the clinic to the laboratory and were immediately processed as previously described[19]. Rhinoprobes were directly transferred into RLT buffer, transported to the lab and processed with the Qiagen RNeasy® Mini Kit according to the manufacturer's instructions.

*MSD Multiplex immunoassay of cytokines and chemokines, ELISA of Tryptase and $LTE_4$*

The thirty-three cytokines and chemokines measured in nMLFs using the electrochemiluminescence technology MSD multiplex U-Plex platform were IL-1α, IL-1β, IL-2, IL-3, IL-4, IL-5, IL-6, IL-7, IL-8, IL-9, IL-10, IL-12p40, IL-12p70, IL-13, IL-15, IL-16, IL-17A, IL-17E/IL-25, IL-21, IL-22, IL-27, IL-33, IL-1RA, IFN-γ, TSLP, TNF-α, TNF-β, GM-CSF, G-CSF, VEGF, CCL11/eotaxin-1, CCL17/TARC, and CCL26/eotaxin-3.

Additionally, tryptase and leukotriene E4 were measured in nMLFs by ELISA. For detailed protocols, upper and lower detection limits and imputation of missing data, please refer to Schneider *et al.*[14]

*Survival analysis*

To assess the cumulative event rate to an ASA reaction in N-ERD patients, Kaplan-Meier plots were used to visualize the survival curves. A log-rank test was applied to compare the curves between the two provocations. R packages *survival* (v3.5-8) and *survminer* (v0.4.9) were employed to compute and visualize survival analysis.

*Hierarchical cluster heatmap*

Thirty-three nasal cytokines and chemokines were log2 transformed and subjected to unsupervised clustering using Euclidean distance metric and ward.D2 linkage method. Cytokines were scaled by row using Z-scores. The correlations were visualised in a heatmap using the R package *pheatmap* (v1.0.12).

*Principal Component Analysis (PCA)*

Principal component analysis of thirty-three scaled raw cytokine and chemokine values was performed at baseline and sixty minutes after reaction onset at provocation one and two in N-ERD patients using the *prcomp* function. Results were visualized with the *factoextra* (v1.0.7) package.

*Cytokine correlations and clustering analysis using CytoMod*

Cytokine-co-clustering was evaluated by the CytoMod software as described[3]. Briefly, cytokines were log10 transformed and clustering was performed using the Pearson-signed metric and complete linkage method. The optimal number of clusters was determined using the Tibshirani gap statistic. Clustering results were visualized by heatmap (*pheatmap*, v1.0.12) with colours denoting the reliability score (yellow: zero reliability, red: high reliability) that was computed across 1,000 random samples and represent the fraction of times each pair of cytokines cluster together. Clustering was performed on adjusted cytokine values.

Module scores for each subject were calculated as the mean value of the standardized (mean zero and unit variance) cytokine values within each module.

*Bulk RNA-Sequencing*

RNA was sequenced using the Lexogen QuantSeq chemistry on an Illumina NextSeq550 system to generate single end reads with 75 bp lengths at the Next Generation Sequencing facility of the Vienna Biocenter (VBCF, Vienna, Austria). To quantify transcripts, the pseudo-mapper *Salmon* was used on a 17 mer index of the human transcriptome (Gencode.v.41)[4].

Quality and adaptor trimming of fastq-files were done with TrimGalore! ( https://github.com/FelixKrueger/TrimGalore/tree/0.6.10) and employing the --*three_prime_clip_R1* argument to clip twelve base pairs from the 3' end of the reads to remove unwanted bias. Mapping was done with the following parameters *-l A, -p 6, --validateMappings, --softclip, --minScoreFraction 0.55.*

After examining the sequences and samples, one sample was excluded because of large sequence duplication rate and one due to potential misclassification. Differential expression analysis was performed with *DESeq2* (v1.42.0)[6]. The *ashr* algorithm was used to shrink the log2-fold changes[7].

*Functional enrichment analysis*

Functional enrichment analyses of significant gene lists were performed using the g:GOSt tool of g:Profiler employing the g:SCS algorithm for calculating significance threshold[8]. A separate analysis was performed for significantly up- and downregulated genes at provocation one and two, respectively.

*Network analysis*

Full STRING networks of DEGs from provocation one and two were constructed using Cytoscape (v3.10.1)[9] with a confidence cut off score of 0.4. Subsequently, networks from provocation one and two were merged and clustered by employing the MCL algorithm with a granularity parameter of 4. Log2 fold changes of both provocations were visualized using Omics Visualizer (v2.0.3).

*Data Availability Statement*

Data has been deposited at the European Genome-phenome Archive (EGA), which is hosted by the EBI and the CRG, under accession number EGAD50000000565. Further information about EGA can be found at https://ega-archive.org and "The European Genome-phenome Archive of human data consented for biomedical research".

#### SUPPLEMENTARY FIGURE LEGENDS

**Supplemental Figure S1. Differences in baseline nasal cytokine levels in disease controls (DC, green, n=10), chronic rhinosinusitis patients with nasal polyps (CRSwNP, purple, n=10) and non-steroidal anti-inflammatory drug (NSAID)-exacerbated respiratory disease (N-ERD, orange, n=11) patients.** (A) – (X) Nasal (A) CCL17, (B) eotaxin-1, (C) GM-CSF (D) IL-1α, (E) IL-1β, (F) IL-2, (G) IL-4, (H) IL-5, (I) IL-6, (J) IL-8, (K) IL-9, (L) IL-10, (M) IL-12p40, (N) IL-13, (O) IL-15, (P) IL-16, (Q) IL-17A, (R) IL-17E/IL-25, (S) IL-33, (T) IFN-γ, (U) TNF-α, (V) VEGF, (W) TSLP, and (X) Tryptase levels at baseline for the three patient groups (x-axes). Boxplots show distribution of absolute cytokine levels (y-axes, pg/mL) with median (horizontal line within box), quartiles (upper and lower box boundaries) and whiskers extending to 1.5 times the inter quartile range. Individual patients are plotted as distinct data points on top. Group-wise comparisons were performed using the Mann-Whitney *U*-test for non-parametric cytokines and tryptase data. Benjamini-Hochberg correction was applied for multiple testing. ns, not significant; *$p < 0.05$; ***$p < 0.001$.

**Supplemental Figure S2. Percentage change in nasal cytokine levels upon ASA provocation in disease controls (DC, green, n=10), chronic rhinosinusitis patients with nasal polyps (CRSwNP, purple, n=10) and non-steroidal anti-inflammatory drug (NSAID)-exacerbated respiratory disease (N-ERD, orange, n=11) patients.**

(A) – (AA) Line graphs showing the median percentage change with median absolute deviation (MAD, y-axes) in (A) IL-4, (B) GM-CSF, (C) IFN-γ, (D) VEGF, (E) IL-6, (F) eotaxin-1, (G) G-CSF, (H) IL-1RA, (I) IL-1α, (J) IL-1β, (K) IL-2, (L) IL-3, (M) IL-7, (N)

IL-8, (O) IL-9, (P) IL-12p70, (Q) IL-13, (R) IL-15, (S) IL-16, (T) IL-17A, (U) IL-17E/IL-25, (V) IL-21, (W) IL-22, (X) IL-27, (Y) IL-33, (Z) TNF-α, and (AA) Tryptase. Timepoint 0 was set at baseline for DC and CRSwNP and at reaction onset for N-ERD patients, respectively.

**Supplemental Figure S3. Changes in nasal mediator levels upon ASA provocation at selected time points in disease controls (DC, green, n=10), chronic rhinosinusitis patients with nasal polyps (CRSwNP, purple, n=10) and non-steroidal anti-inflammatory drug (NSAID)-exacerbated respiratory disease (N-ERD, orange, n=11) patients.** (A) – (AA) Boxplots showing absolute cytokine levels (y-axes, pg/mL) of (A) IL-4, (B) GM-CSF, (C) IFN-γ, (D) VEGF, (E) IL-6, (F) eotaxin-1, (G) G-CSF, (H) IL-1RA, (I) IL-1α, (J) IL-1β, (K) IL-2, (L) IL-3, (M) IL-7, (N) IL-8, (O) IL-9, (P) IL-12p70, (Q) IL-13, (R) IL-15, (S) IL-16, (T) IL-17A, (U) IL-17E/IL-25, (V) IL-21, (W) IL-22, (X) IL-27, (Y) IL-33, (Z) TNF-α, and (AA) Tryptase at indicated timepoints after ASA provocation in the three patient groups (x-axes). Timepoint 0 was set at baseline for DC and CRSwNP and at reaction onset for N-ERD patients, respectively. Group-wise comparisons were performed using the Mann-Whitney *U*-test with Benjamini-Hochberg correction. *$p < 0.05$; **$p < 0.01$; ***$p < 0.001$; ****$p < 0.0001$; non-significant not shown.

**Supplemental Figure S4. Changes in nasal mediator levels upon ASA provocation before (n=10, blue) and after twenty-four weeks (n=10, fuchsia) of dupilumab treatment in non-steroidal anti-inflammatory drug (NSAID)-exacerbated disease (N-ERD) patients.**

(A) – (AA) Changes in (A) VEGF, (B) G-CSF, (C) GM-CSF, (D) eotaxin-1, (E) IL-1α, (F) IL-1β, (G) IL-1RA, (H) IL-2, (I) IL-3, (J) IL-4, (K) IL-6, (L) IL-7, (M) IL-8, (N) IL-9, (O) IL-12p70, (P) IL-13, (Q) IL-15, (R) IL-16, (S) IL-17A, (T) IL-17E/IL-25, (U) IL-21, (V) IL-22, (W) IL-27, (X) IL-33, (Y) IFN-γ, (Z) TNF-α, and (AA) Tryptase levels in N-ERD patients. Barplots show median values for area under the curve (AUC) for the cytokines from time point of reaction onset to sixty minutes post treatment of provocation 1 and 2 as indicated. Error bars denote 95% confidence intervals. Differences were tested using the Wilcoxon signed-rank test. ns, not significant; $*p < 0.05$; $**p < 0.01$.

**Supplemental Figure S5. Changes in nasal mediator levels upon ASA provocation in non-steroidal anti-inflammatory drug (NSAID)-exacerbated disease (N-ERD, n=11) patients at provocation 1 (blue) and 2 (fuchsia).**

(A) – (AI) Changes in (A) $LTE_4$, (B) TSLP, (C) IL-5, (D) CCL17, (E) eotaxin-3, (F) IL-10, (G) IL-12p40, (H) TNF-β, (I) eotaxin-1, (J) G-CSF, (K) GM-CSF, (L) IFN-γ , (M) IL-1RA, (N) IL-1α, (O) IL-1β, (P) IL-2, (Q) IL-3, (R) IL-4, (S) IL-6, (T) IL-7, (U) IL-8, (V) IL-9, (W) IL-12p70, (X) IL-13, (Y) IL-15, (Z) IL-16, (AA) IL-17A, (AB) IL17E/IL-25, (AC) IL-21, (AD) IL-22, (AE) IL-27, (AF) IL-33, (AG) TNF-α, (AH) VEGF, and (AI) Tryptase. Panels depict absolute mediator levels (y-axes, pg/mL) in individual patients at provocation 1 from baseline (start of provocation) across full treatment period.

**Supplemental Figure S6. Principal component and clustering analysis of nasal mediator levels in non-steroidal anti-inflammatory drug (NSAID)-exacerbated respiratory disease before (n=11 baseline and n=10 post 60) and twenty-four weeks after (n=11 baseline and n=10 post 60) dupilumab therapy.**

(A) Principal component analysis of nasal mediator levels at provocation 1 baseline, visualising principal components 1 and 2 (PC1 and PC2). Colour and intensity of arrows, corresponding to one of the thirty-three mediators, indicate the strength and direction of their contribution to the respective principal component.

(B) Distribution of N-ERD subjects, grouped into ASA-tolerant (red) and ASA-intolerant (orange), within PC1 and PC2.

(C) – (F) Heatmaps with cluster membership and reliability scores of adjusted cytokines and module scores comparing ASA-tolerant and intolerant patients (D) before (provocation 1) and (F) twenty-four weeks (provocation 2) after start of dupilumab treatment.

**Supplemental Figure S7. Changes in nasal mediator levels in anti-inflammatory drug (NSAID)-exacerbated disease (N-ERD) patients at baseline (ASA-intolerant: n=5, yellow and ASA-tolerant: n=5, red) and after twenty-four weeks (ASA-intolerant: n=6, yellow and ASA-tolerant: n=4, red) of dupilumab treatment.**

(A) – (AI) Changes in (A) CCL17, (B) eotaxin-1, (C) eotaxin-3, (D) G-CSF, (E) GM-CSF, (F) IFN-γ, (G) IL-1RA, (H) IL-1α, (I) IL-1β, (J) IL-2, (K) IL-3, (L) IL-4, (M) IL-5, (N) IL-6, (O) IL-7, (P) IL-8, (Q) IL-9, (R) IL-10, (S) IL-12p40, (T) IL-12p70, (U) IL-13, (V) IL-15, (W) IL-16, (X) IL-17A, (Y) IL-17E/IL-25, (Z) IL-21, (AA) IL-22, (AB) IL-27, (AC) IL-33, (AD) TNF-α, (AE) TNF-β, (AF) TSLP, (AG) VEGF, (AH) Tryptase and, (AI) $LTE_4$. Paired boxplots show distribution of absolute cytokine levels (y-axes, pg/mL) with median (horizontal line within box), quartiles (upper and lower box boundaries) and whiskers extending to 1.5 times the interquartile range at reaction onset and sixty minutes thereafter at provocation 1 and 2. Individual patients are plotted as distinct data points on top. Pair-wise comparisons were performed using the Wilcoxon signed rank test.

**Supplemental Figure S8. Nasal transcriptome signature of non-steroidal anti-inflammatory drug (NSAID)-exacerbated respiratory disease patients (N-ERD) before (n=11 before provocation 1 and n=10 after provocation 1) and twenty-four weeks after (n=10 before provocation 2 and n=11 after provocation 2) dupilumab therapy.**

(A) – (D) Boxplots showing the relative expression (y-axes, log2) of (A) *ALOX15*, (B) *APOE*, (C) *AREG*, and (D) *NOS2* with median (horizontal line within box), quartiles (upper and lower box boundaries) and whiskers extending to 1.5 times the interquartile range before and after ASA challenge at provocation 1 (P1) and 2 (P2) for ASA-intolerant (circles) and tolerant (triangles) groups. Individual patients are plotted as distinct data points on top. Group-wise comparisons were done using a *t*-test. ns, not significant; *$p < 0.05$; **$p < 0.01$; ***$p < 0.001$.

#### SUPPLEMENTARY TABLES

**Table S1. Lower and upper respiratory tract reactions in N-ERD patients before and sixty minutes after ASA provocation at baseline and after twenty-four weeks of dupilumab therapy.**

| | **Provocation 1** *N* = 11[1] | | **Provocation 2** *N* = 11[1] | | |
|---|---|---|---|---|---|
| **Respiratory variable** | **Before** | **60 min after** | **Before** | **60 min after** | ***p*-value**[2] |
| **LRT** | 0 (0%) | 8 (80%) | 0 (0%) | 0 (0%) | <0.001 |
| *Unknown* | 0 | 1 | 0 | 1 | |
| **URT** | 3 (27%) | 8 (80%) | 0 (0%) | 8 (80%) | <0.001 |
| *Unknown* | 0 | 1 | 0 | 1 | |
| **$FEV_1$** | 290 (264, 353) | 200 (160, 258) | 340 (323, 397) | 313 (305, 380) | 0.002 |
| *Unknown* | 0 | 0 | 0 | 1 | |

[1]n (%); Median (inter quartile range, IQR)

[2]Fisher's exact test; Kruskal-Wallis rank sum test

LRT: Lower respiratory tract reaction; URT: Upper respiratory tract reaction

$FEV_1$: Forced expiratory volume in one minute

**Table S2. Selected results from functional enrichment analysis of differentially expressed genes (DEGs) at provocation 1 and 2.**

| **Provocation 1 – upregulated DEGs** | | | | | |
|---|---|---|---|---|---|
| **Source** | **Term name** | **Term ID** | **adj. *p*-value** | **Intersection size** | **Genes** |
| GO:MF | Cytokine activity | GO:0005125 | $7.26 \times 10^{-3}$ | 22 | *NRG1, CLCF1, IL1RN, GDF7, BMP5, CMTM7, IL37, BMP8A, SLURP1, TNFSF18, CCL22, IL36RN, AREG, INHBE, EDN1, BMP6, IL36G, WNT9B, ADIPOQ, CCL16, IL5, IL11* |
| GO:BP | Inflammatory response | GO:0006954 | $4.10 \times 10^{-2}$ | 49 | *ANXA1, EPHA2, C5AR2, F12, LILRA5, HAMP, IL1RN, SERPINE1, THBS1, GGT5, HAVCR2, OLR1, S100A8, ADM, NLRC3, IRGM, AGT, LRRC19, IL37, BTK, TRPV1, IL1RL1, SLC11A1, SEMA7A, CCR4, ECM1, TEK, GPR4, TNFSF18, KRT16, PTGIS, TSPAN18, CCL22, IL36RN, FABP4, PIK3CD, BMP6, SCUBE1, MEFV, IL36G, SPINK7, PLA2G4B, ADIPOQ, WNK4, PLD4, AZU1, CCL16, IL5, TBXA2R* |

| **Provocation 2 – upregulated DEGs** | | | | | |
|---|---|---|---|---|---|
| **Source** | **Term name** | **Term ID** | **adj. *p*-value** | **Intersection size** | **Genes** |
| KEGG | Arachidonic acid metabolism | KEGG:00590 | $3.41 \times 10^{-2}$ | 14 | *CYP2C19, PLA2G4B, PTGIS, PLA2G2F, PLA2G4D, PLA2G4E, PLA2G4C, ALOX12B, GGT5, CYP2C9, CYP4F3, ALOXE3, CYP2E1, ALOX15B* |
| GO:MF | RAGE receptor binding | GO:0050786 | $1.01 \times 10^{-2}$ | 6 | *S100A7, S100A8, S100A9, FPR1, S100A12, S100B* |

| **Provocation 2 – downregulated DEGs** | | | | | |
|---|---|---|---|---|---|
| **Source** | **Term name** | **Term ID** | **adj. *p*-value** | **Intersection size** | **Genes** |
| KEGG | Peroxisome | KEGG:04146 | $5{,}72 \times 10^{-7}$ | 28 | *NOS2, ABCD2, ACOT8, SLC27A2, ECI2, NUDT7, PEX14, PRDX5, PEX12, PEX10, PEX6, PEX16, PEX3, CAT, HACL1, DDO, ABCD3, PEX2, PEX19, SOD1, PHYH, GNPAT, SCP2, ACSL3, PEX7, NUDT12, SLC25A17, MPV17* |

KEGG: Kyoto Encyclopedia of Genes and Genomes
GO:MF: Gene Ontology Molecular Function
GO:BP: Gene Ontology Biological Process

Figure S1

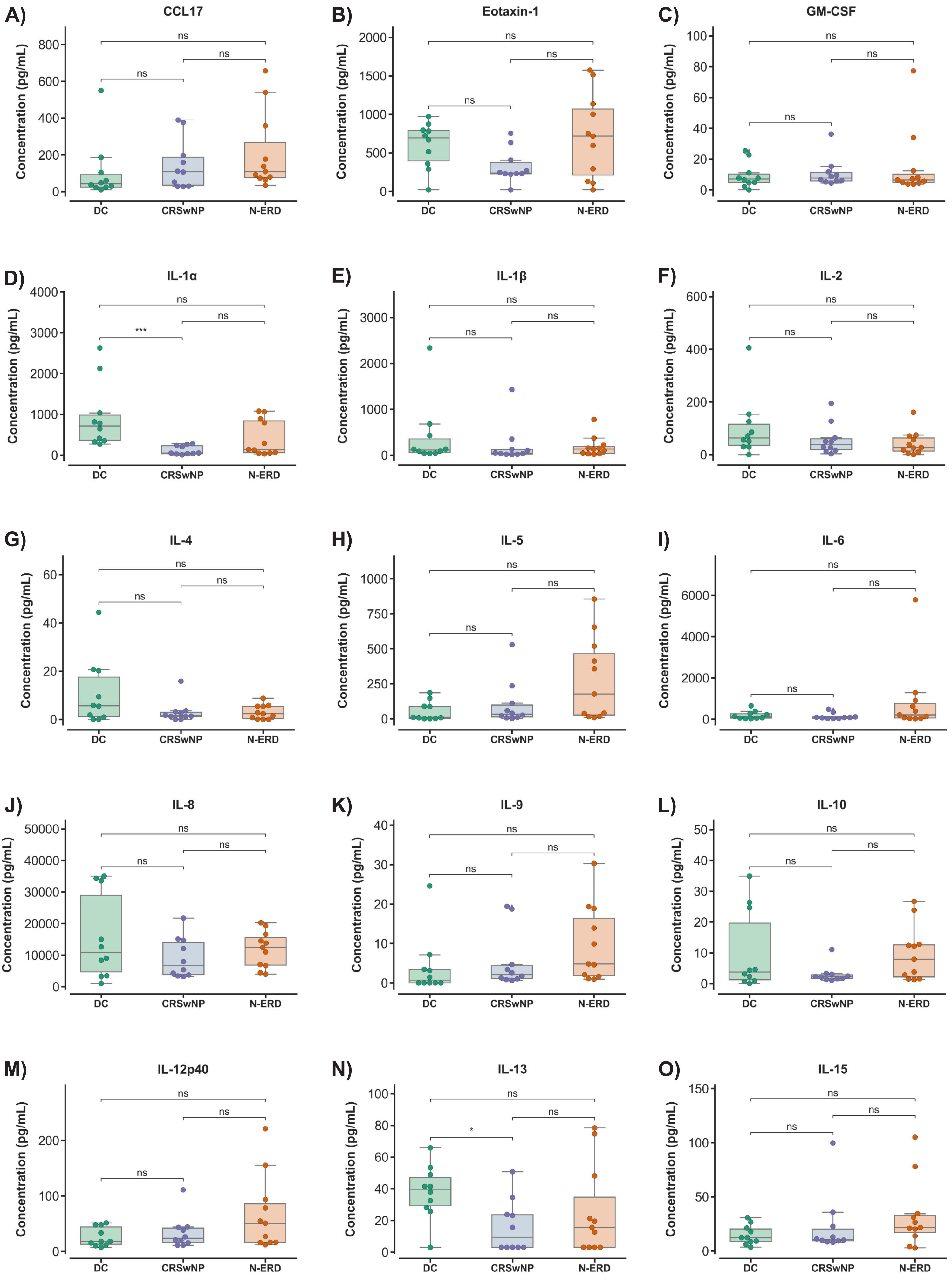
A) CCL17
B) Eotaxin-1
C) GM-CSF
D) IL-1α
E) IL-1β
F) IL-2
G) IL-4
H) IL-5
I) IL-6
J) IL-8
K) IL-9
L) IL-10
M) IL-12p40
N) IL-13
O) IL-15
Concentration (pg/mL)
DC
CRSwNP
N-ERD
ns
***
*

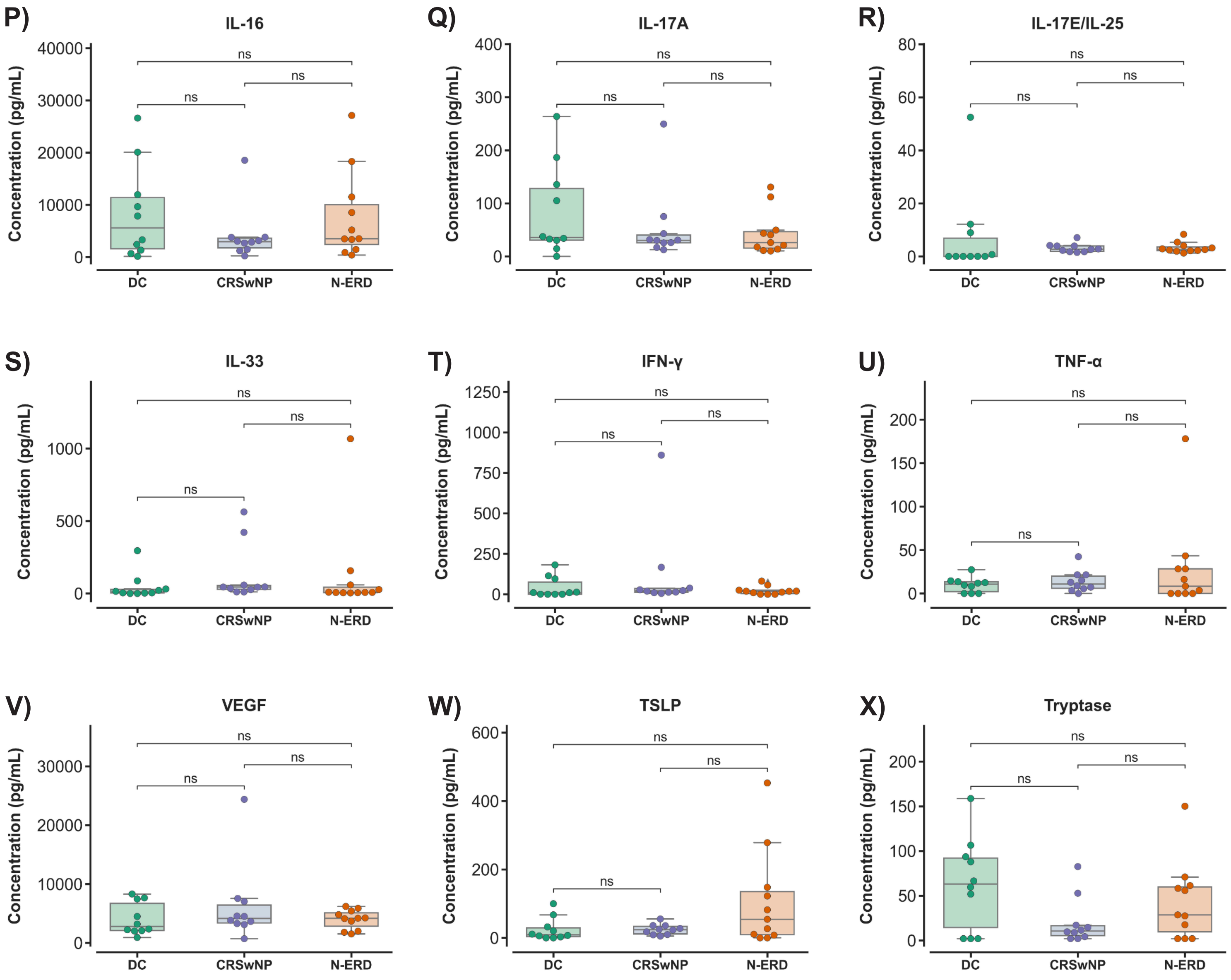

P)
IL-16
Q)
IL-17A
R)
IL-17E/IL-25
S)
IL-33
T)
IFN-γ
U)
TNF-α
V)
VEGF
W)
TSLP
X)
Tryptase
Concentration (pg/mL)
DC
CRSwNP
N-ERD
ns

Figure S2

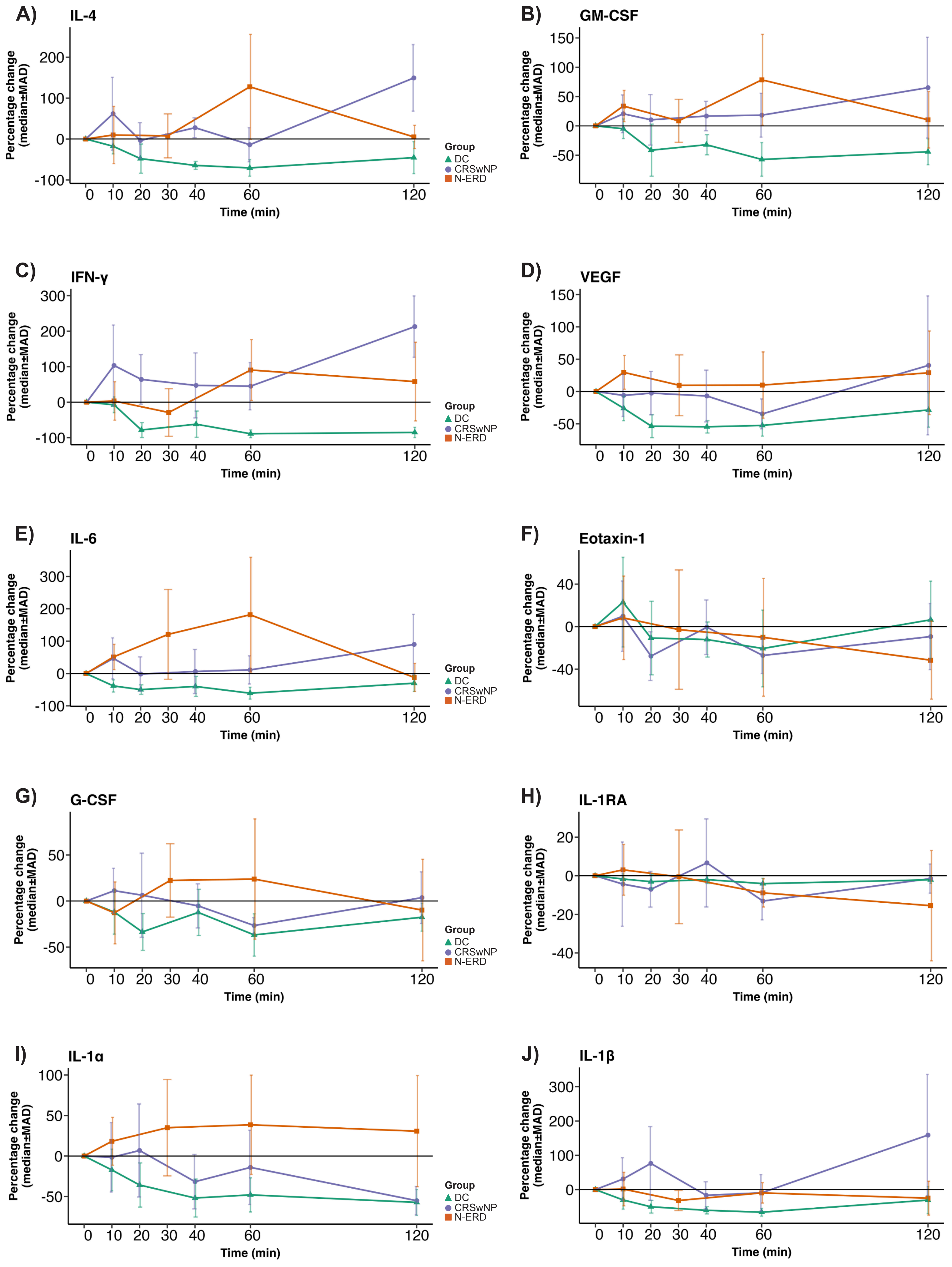

A)
IL-4
B)
GM-CSF
C)
IFN-γ
D)
VEGF
E)
IL-6
F)
Eotaxin-1
G)
G-CSF
H)
IL-1RA
I)
IL-1α
J)
IL-1β
Percentage change (median±MAD)
Time (min)
Group
DC
CRSwNP
N-ERD

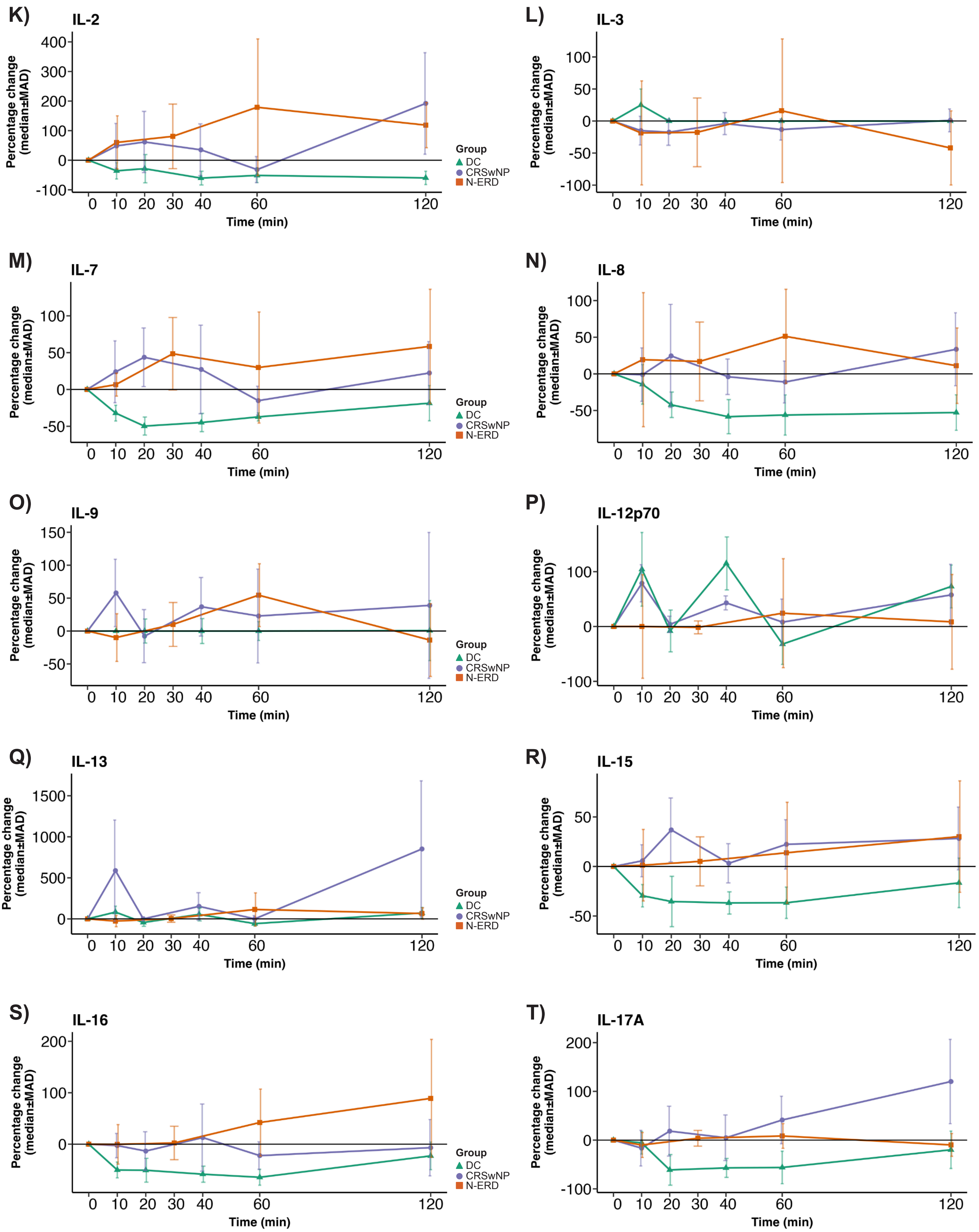
K)
IL-2
Percentage change (median±MAD)
400
300
200
100
0
-100
0 10 20 30 40 60 120
Time (min)
Group
DC
CRSwNP
N-ERD
L)
IL-3
Percentage change (median±MAD)
100
50
0
-50
-100
0 10 20 30 40 60 120
Time (min)
M)
IL-7
Percentage change (median±MAD)
100
50
0
-50
0 10 20 30 40 60 120
Time (min)
Group
DC
CRSwNP
N-ERD
N)
IL-8
Percentage change (median±MAD)
100
50
0
-50
0 10 20 30 40 60 120
Time (min)
O)
IL-9
Percentage change (median±MAD)
150
100
50
0
-50
0 10 20 30 40 60 120
Time (min)
Group
DC
CRSwNP
N-ERD
P)
IL-12p70
Percentage change (median±MAD)
100
0
-100
0 10 20 30 40 60 120
Time (min)
Q)
IL-13
Percentage change (median±MAD)
1500
1000
500
0
0 10 20 30 40 60 120
Time (min)
Group
DC
CRSwNP
N-ERD
R)
IL-15
Percentage change (median±MAD)
50
0
-50
0 10 20 30 40 60 120
Time (min)
S)
IL-16
Percentage change (median±MAD)
200
100
0
0 10 20 30 40 60 120
Time (min)
Group
DC
CRSwNP
N-ERD
T)
IL-17A
Percentage change (median±MAD)
200
100
0
-100
0 10 20 30 40 60 120
Time (min)

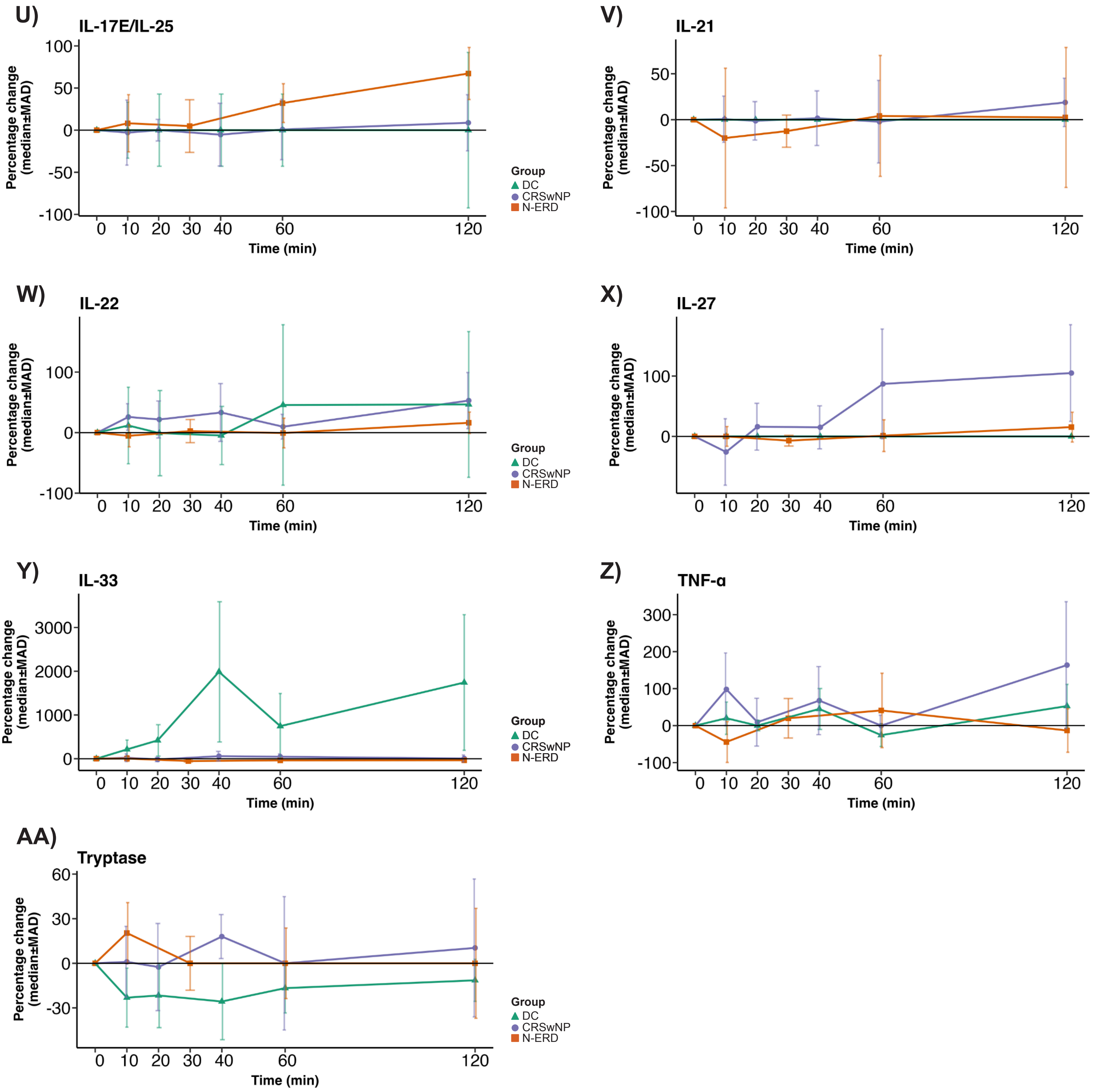

U)
IL-17E/IL-25
Percentage change (median±MAD)
100
50
0
-50
-100
0 10 20 30 40 60 120
Time (min)
Group
DC
CRSwNP
N-ERD
V)
IL-21
Percentage change (median±MAD)
50
0
-50
-100
0 10 20 30 40 60 120
Time (min)
W)
IL-22
Percentage change (median±MAD)
100
0
-100
0 10 20 30 40 60 120
Time (min)
Group
DC
CRSwNP
N-ERD
X)
IL-27
Percentage change (median±MAD)
100
0
0 10 20 30 40 60 120
Time (min)
Y)
IL-33
Percentage change (median±MAD)
3000
2000
1000
0
0 10 20 30 40 60 120
Time (min)
Group
DC
CRSwNP
N-ERD
Z)
TNF-α
Percentage change (median±MAD)
300
200
100
0
-100
0 10 20 30 40 60 120
Time (min)
AA)
Tryptase
Percentage change (median±MAD)
60
30
0
-30
0 10 20 30 40 60 120
Time (min)
Group
DC
CRSwNP
N-ERD

Figure S3

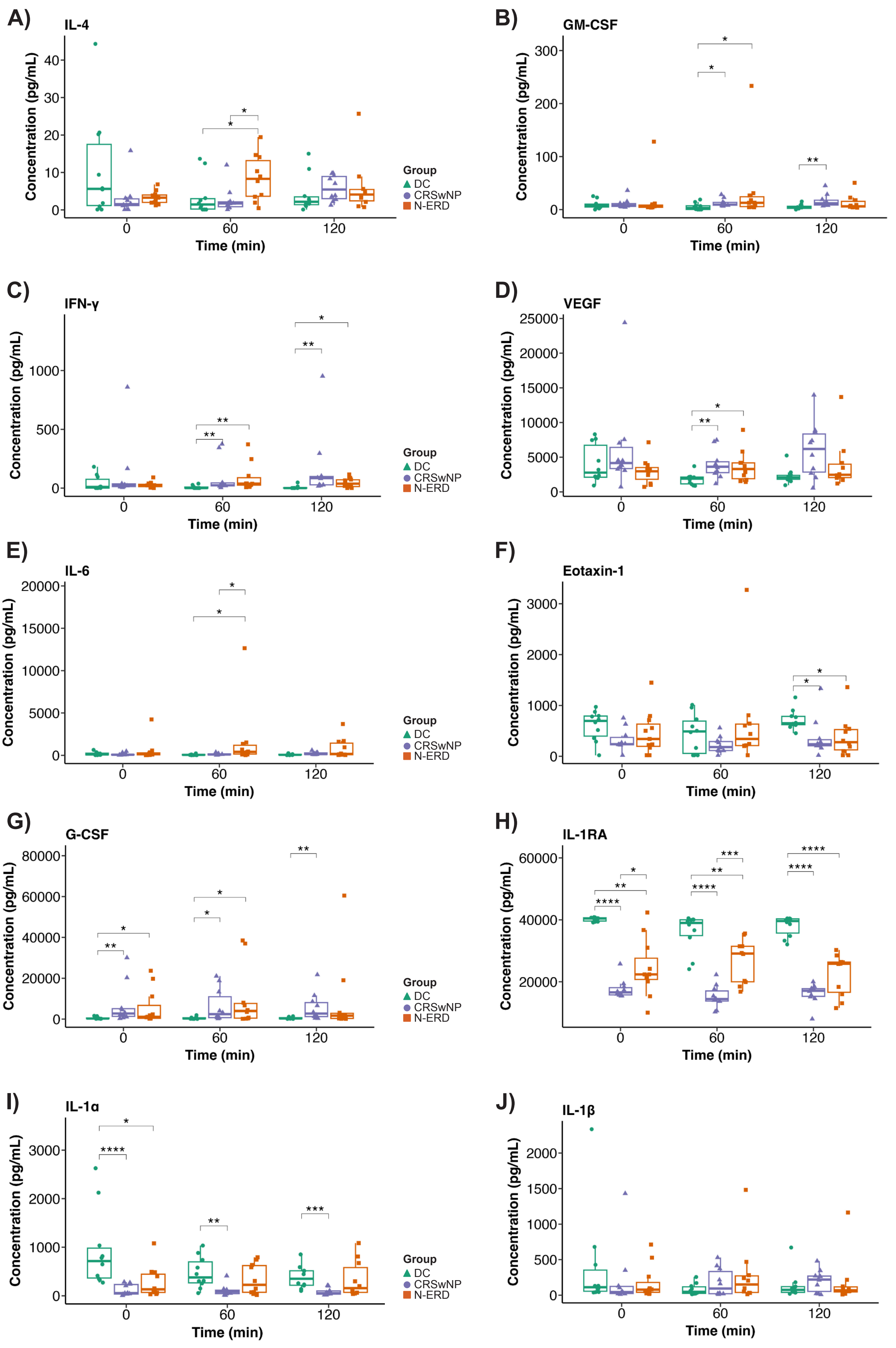
A)
IL-4
B)
GM-CSF
C)
IFN-γ
D)
VEGF
E)
IL-6
F)
Eotaxin-1
G)
G-CSF
H)
IL-1RA
I)
IL-1α
J)
IL-1β
Concentration (pg/mL)
Time (min)
0
60
120
Group
DC
CRSwNP
N-ERD

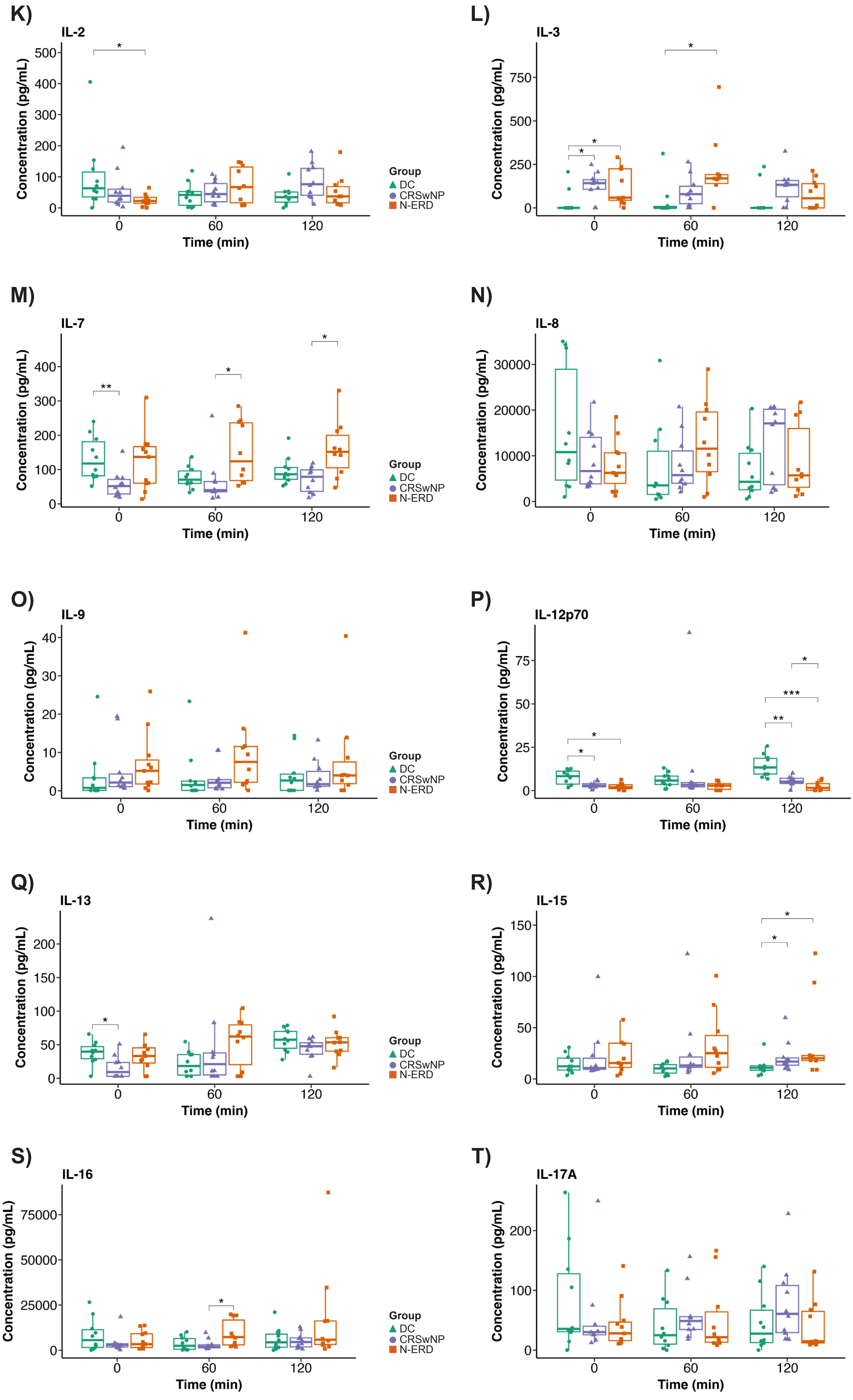
K)
IL-2
L)
IL-3
M)
IL-7
N)
IL-8
O)
IL-9
P)
IL-12p70
Q)
IL-13
R)
IL-15
S)
IL-16
T)
IL-17A
Concentration (pg/mL)
Time (min)
0
60
120
Group
DC
CRSwNP
N-ERD
*
**
***

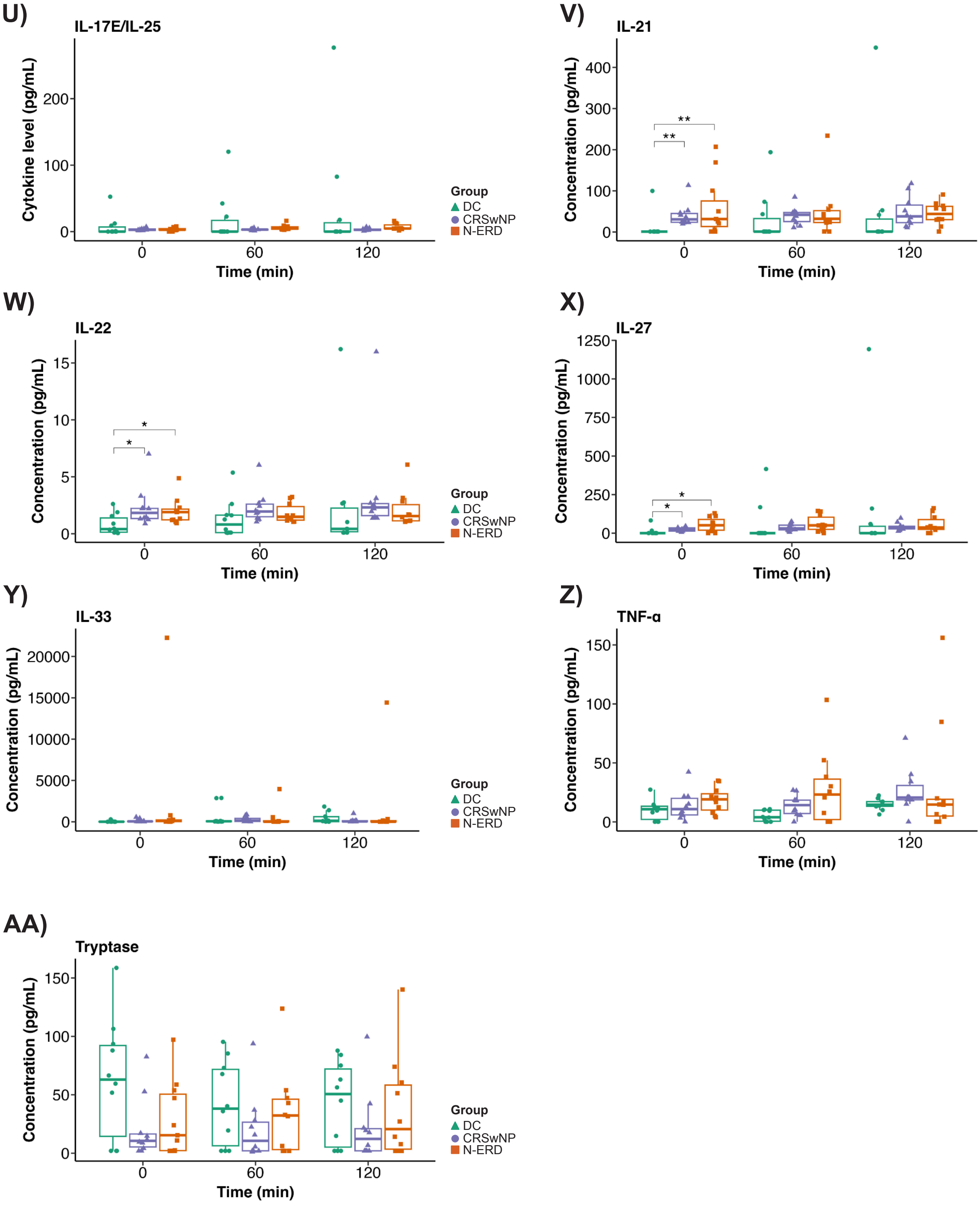
U)
IL-17E/IL-25
Cytokine level (pg/mL)
Time (min)
V)
IL-21
Concentration (pg/mL)
Time (min)
W)
IL-22
Concentration (pg/mL)
Time (min)
X)
IL-27
Concentration (pg/mL)
Time (min)
Y)
IL-33
Concentration (pg/mL)
Time (min)
Z)
TNF-α
Concentration (pg/mL)
Time (min)
AA)
Tryptase
Concentration (pg/mL)
Time (min)
Group
DC
CRSwNP
N-ERD

### Figure S4

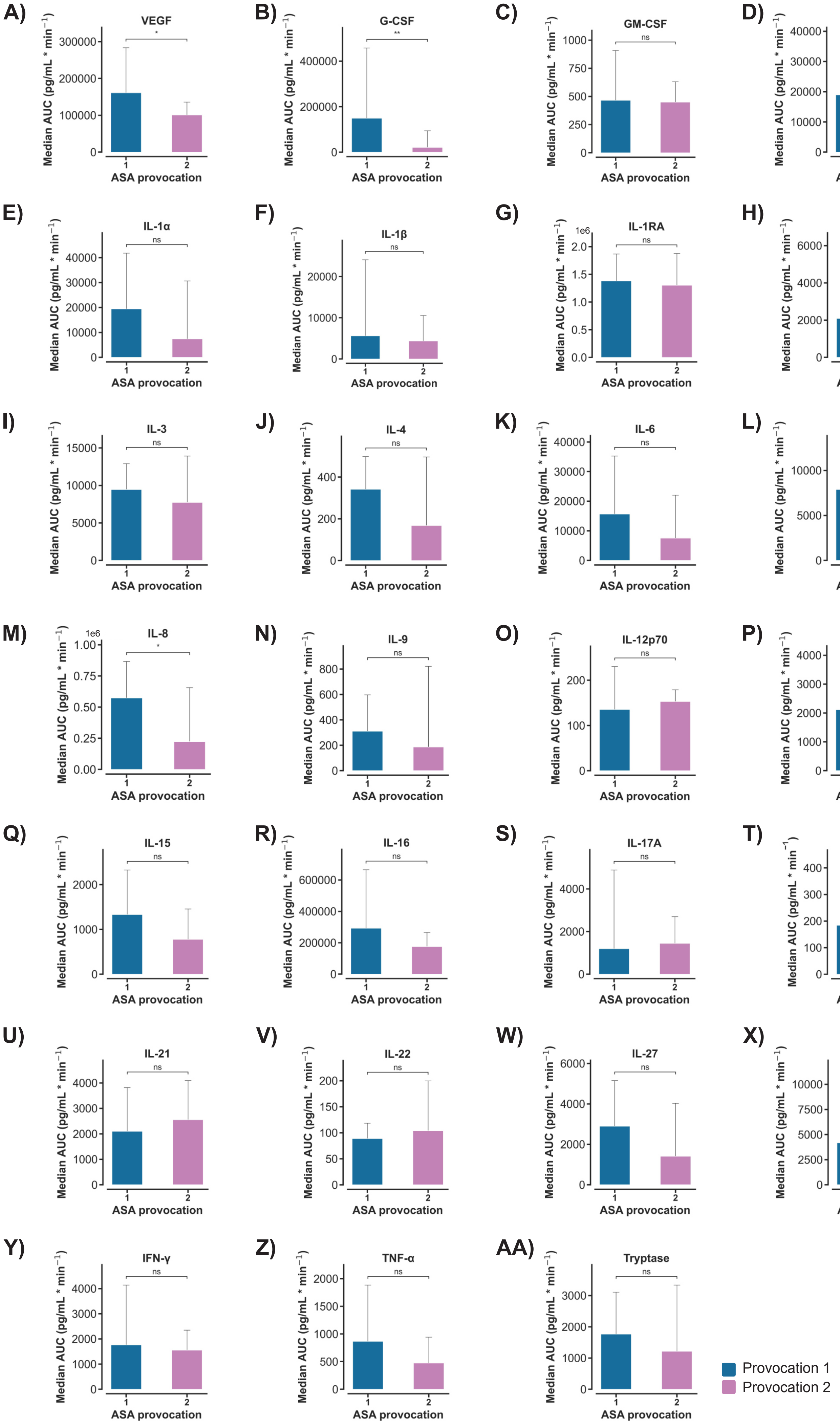

A) VEGF *
B) G-CSF **
C) GM-CSF ns
D) Eotaxin-1 ns
E) IL-1α ns
F) IL-1β ns
G) IL-1RA ns
1e6
H) IL-2 ns
I) IL-3 ns
J) IL-4 ns
K) IL-6 ns
L) IL-7 ns
M) IL-8 *
1e6
N) IL-9 ns
O) IL-12p70 ns
P) IL-13 ns
Q) IL-15 ns
R) IL-16 ns
S) IL-17A ns
T) IL-17E/IL-25 ns
U) IL-21 ns
V) IL-22 ns
W) IL-27 ns
X) IL-33 ns
Y) IFN-γ ns
Z) TNF-α ns
AA) Tryptase ns
Median AUC (pg/mL * min$^{-1}$)
ASA provocation
1
2
Provocation 1
Provocation 2

Figure S5

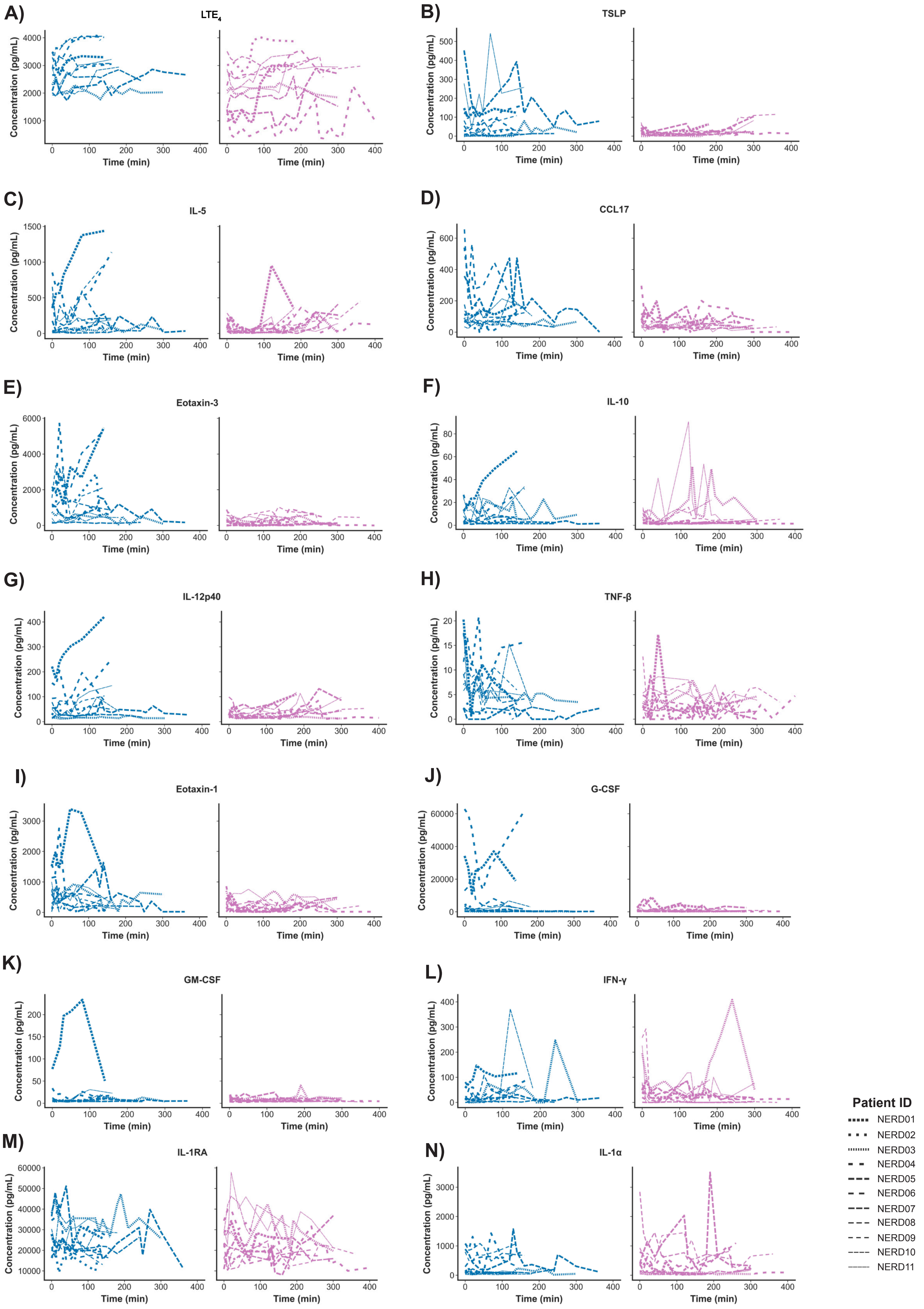

A)
LTE4
B)
TSLP
C)
IL-5
D)
CCL17
E)
Eotaxin-3
F)
IL-10
G)
IL-12p40
H)
TNF-β
I)
Eotaxin-1
J)
G-CSF
K)
GM-CSF
L)
IFN-γ
M)
IL-1RA
N)
IL-1α
Concentration (pg/mL)
Time (min)
Patient ID
NERD01
NERD02
NERD03
NERD04
NERD05
NERD06
NERD07
NERD08
NERD09
NERD10
NERD11

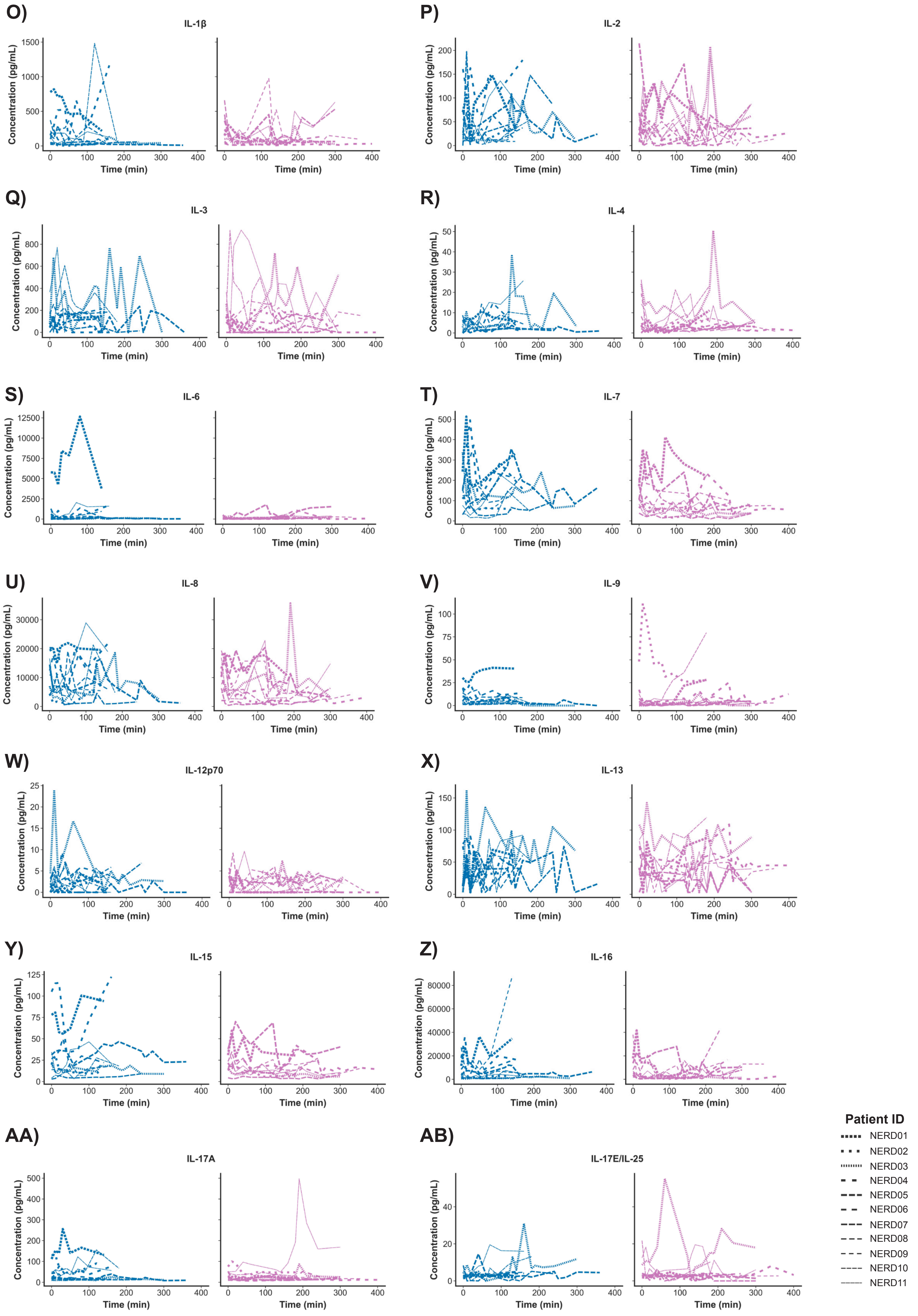
O)
IL-1β
Concentration (pg/mL)
Time (min)
P)
IL-2
Q)
IL-3
R)
IL-4
S)
IL-6
T)
IL-7
U)
IL-8
V)
IL-9
W)
IL-12p70
X)
IL-13
Y)
IL-15
Z)
IL-16
AA)
IL-17A
AB)
IL-17E/IL-25
Patient ID
NERD01
NERD02
NERD03
NERD04
NERD05
NERD06
NERD07
NERD08
NERD09
NERD10
NERD11

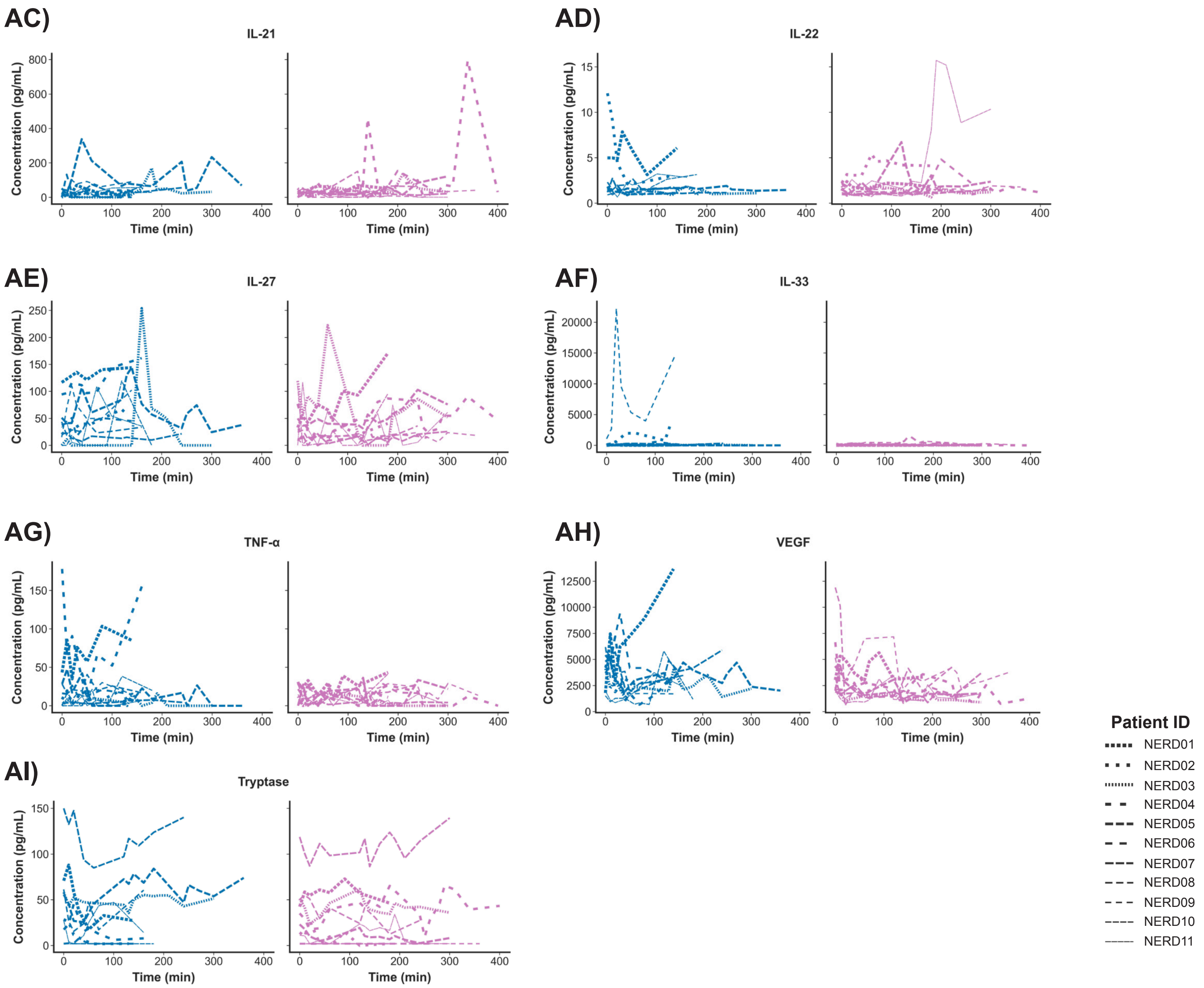
AC)
IL-21
AD)
IL-22
AE)
IL-27
AF)
IL-33
AG)
TNF-α
AH)
VEGF
AI)
Tryptase
Concentration (pg/mL)
Time (min)
Patient ID
NERD01
NERD02
NERD03
NERD04
NERD05
NERD06
NERD07
NERD08
NERD09
NERD10
NERD11

Figure S6

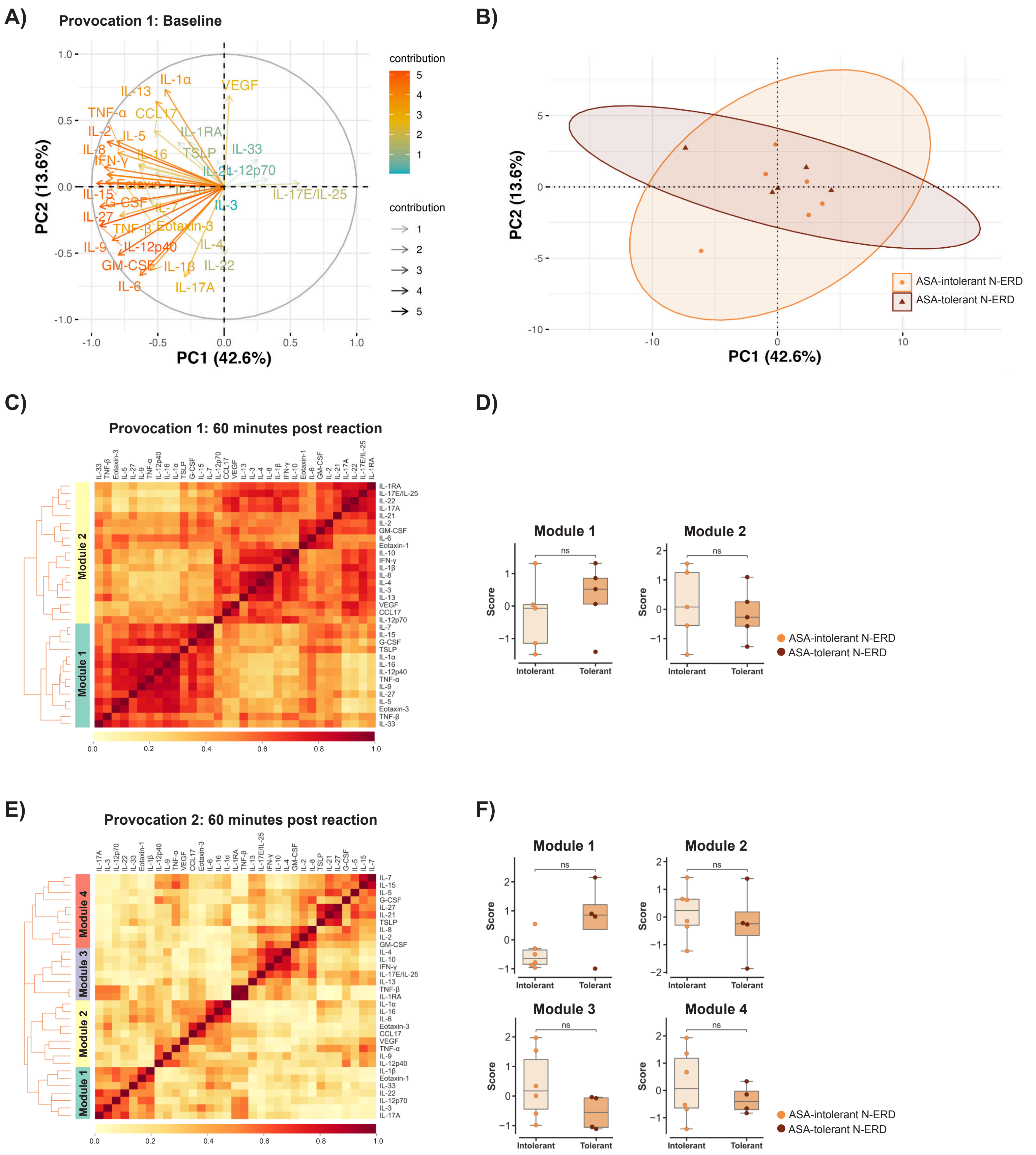

A)
Provocation 1: Baseline
PC1 (42.6%)
PC2 (13.6%)
contribution
B)
ASA-intolerant N-ERD
ASA-tolerant N-ERD
C)
Provocation 1: 60 minutes post reaction
Module 1
Module 2
D)
Module 1
Module 2
Score
Intolerant
Tolerant
ns
ASA-intolerant N-ERD
ASA-tolerant N-ERD
E)
Provocation 2: 60 minutes post reaction
Module 1
Module 2
Module 3
Module 4
F)
Module 1
Module 2
Module 3
Module 4
Score
Intolerant
Tolerant
ns
ASA-intolerant N-ERD
ASA-tolerant N-ERD

Figure S7

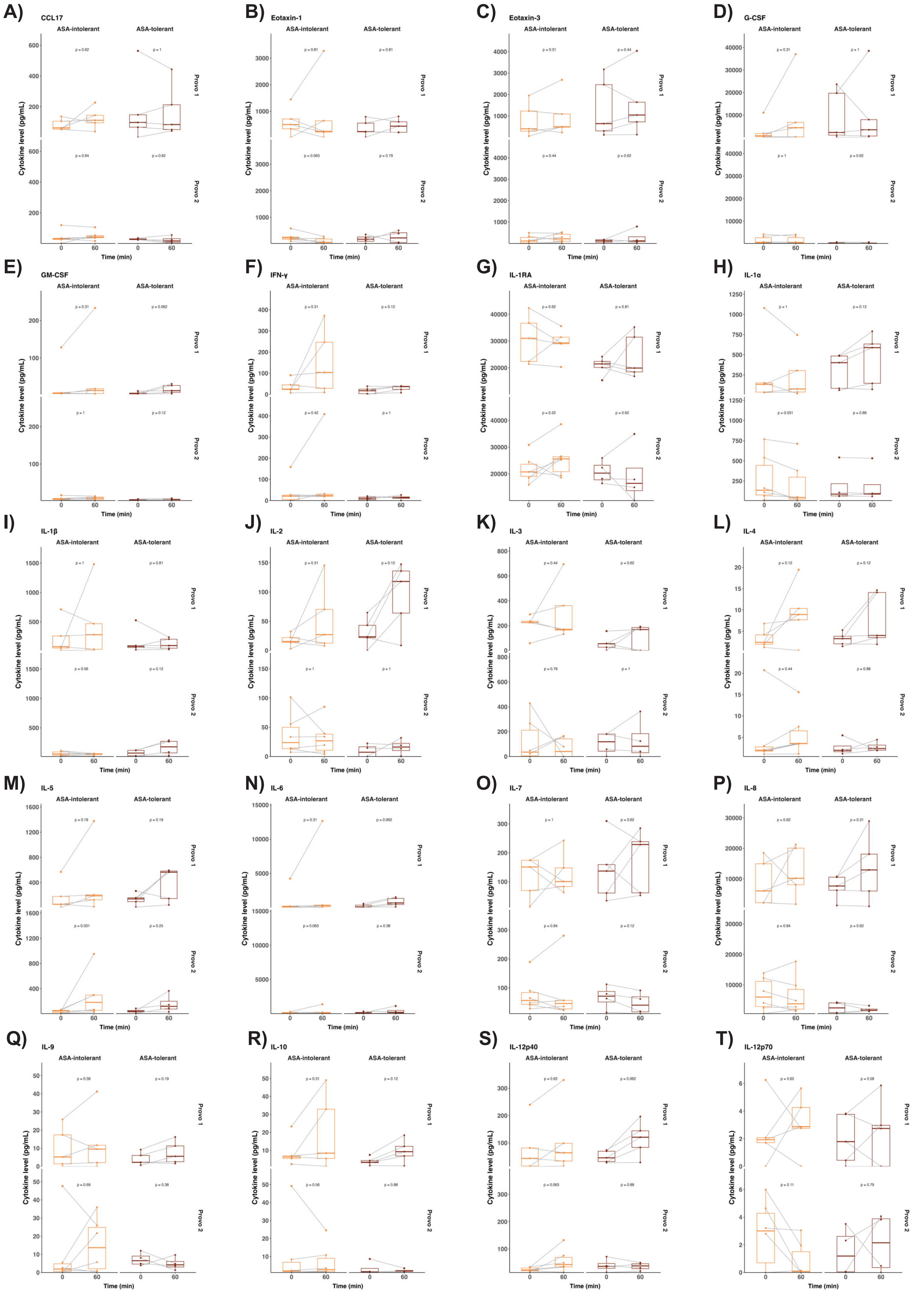

A) CCL17
B) Eotaxin-1
C) Eotaxin-3
D) G-CSF
E) GM-CSF
F) IFN-γ
G) IL-1RA
H) IL-1α
I) IL-1β
J) IL-2
K) IL-3
L) IL-4
M) IL-5
N) IL-6
O) IL-7
P) IL-8
Q) IL-9
R) IL-10
S) IL-12p40
T) IL-12p70
ASA-intolerant
ASA-tolerant
Provo 1
Provo 2
Cytokine level (pg/mL)
Time (min)

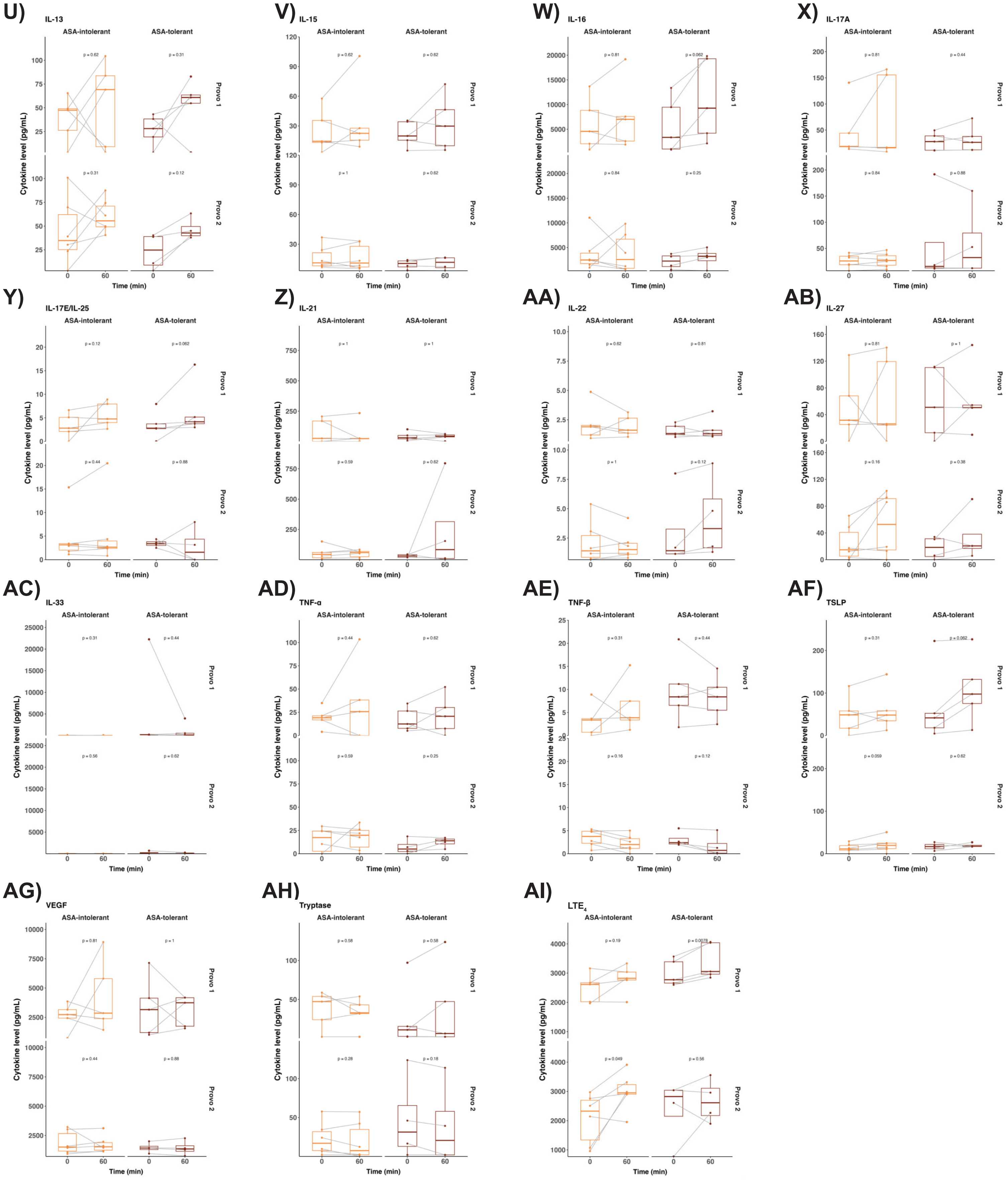

U)
IL-13
V)
IL-15
W)
IL-16
X)
IL-17A
Y)
IL-17E/IL-25
Z)
IL-21
AA)
IL-22
AB)
IL-27
AC)
IL-33
AD)
TNF-α
AE)
TNF-β
AF)
TSLP
AG)
VEGF
AH)
Tryptase
AI)
LTE4
ASA-intolerant
ASA-tolerant
Provo 1
Provo 2
Cytokine level (pg/mL)
Time (min)

### Figure S8

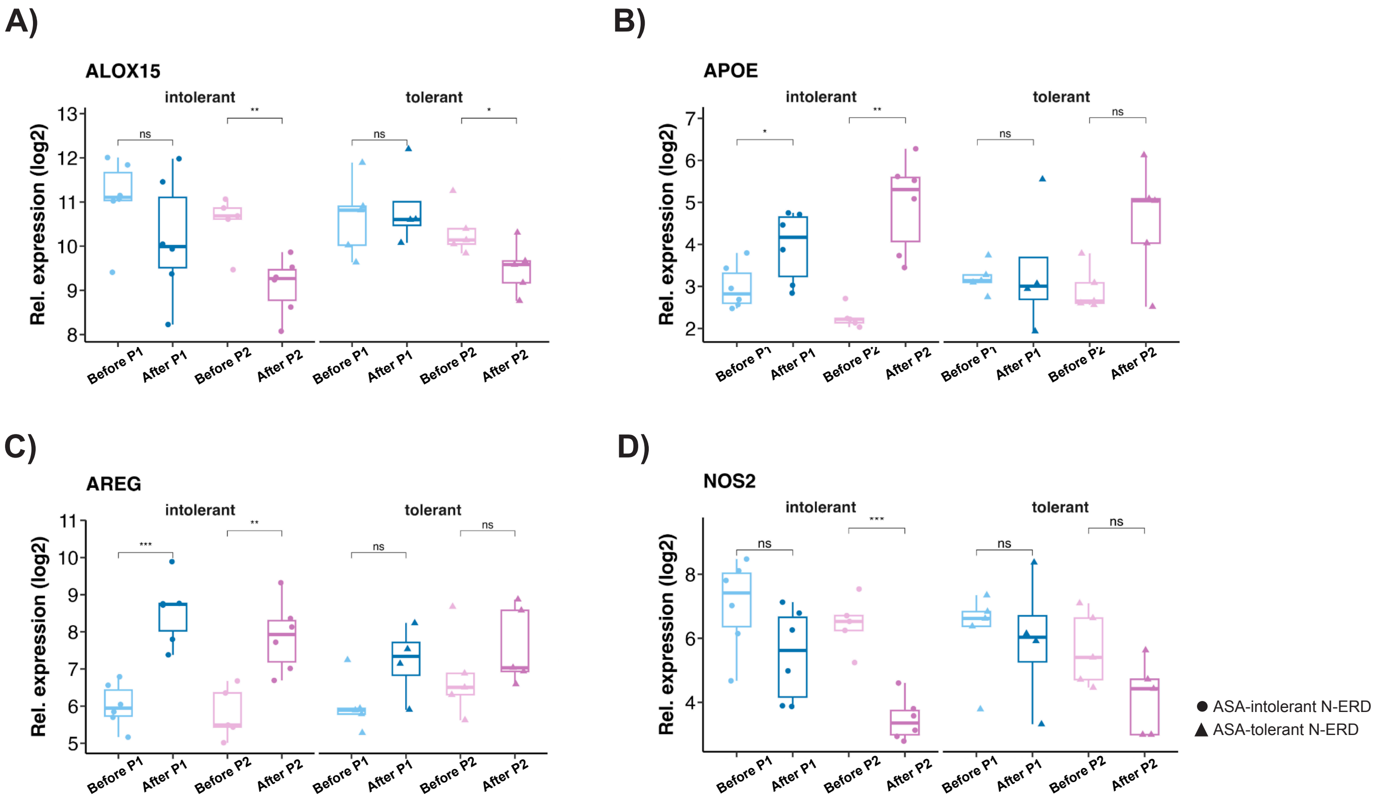
A)
ALOX15
intolerant
tolerant
Rel. expression (log2)
Before P1
After P1
Before P2
After P2
B)
APOE
C)
AREG
D)
NOS2
ASA-intolerant N-ERD
ASA-tolerant N-ERD
